# Comparative Efficacy and Safety of Novel Oral Anticoagulants in Atrial Fibrillation with Chronic Kidney Disease: A Systematic Review, Meta-Regression, and Network Meta-Analysis

**DOI:** 10.1101/2025.08.28.25334228

**Authors:** Manish Juneja, Rakhshanda Khan, Gaurav Sharma, Nitish Ajit Nadkarni, Arnav Gandhi, Chidurala Rahul, Mohd Sijad Uddin, Samriddhi Sharma, Ambala Madhulikha, Bharadwaj Jilakraju, Syeda Hafsa Noor Ain, Harshawardhan Dhanraj Ramteke

## Abstract

**Background:** Patients with both atrial fibrillation (AF) and chronic kidney disease (CKD) face a difficult clinical trade-off: they need protection from stroke but are at higher risk of bleeding. With more anticoagulant options now available, especially among newer oral agents, it’s unclear which offers the safest and most effective choice in this high-risk group.

**Methods:** We conducted a systematic review and network meta-analysis of randomized controlled trials involving adults with AF and CKD. We searched five major databases, focusing on three key outcomes: all-cause mortality, stroke or systemic embolism, and significant bleeding. Analyses were performed using MetaInsight in R, and risk of bias was assessed using Cochrane’s RoB 2.0 tool.

**Results:** We included 29 trials with over 187,000 participants. Apixaban stood out across outcomes, showing the lowest risk of all-cause mortality (OR 0.45; 95% CI: 0.22–0.92) and stroke/systemic embolism (OR 0.39; 95% CI: 0.25–0.62), along with a favorable bleeding profile. Asundexian, a newer factor XIa inhibitor, showed some early promise in lowering stroke risk, but there’s still not enough evidence to know how safe it really is. In contrast, warfarin and aspirin repeatedly came up short, providing less protection and carrying more risk than the newer options. These patterns remained consistent across all sensitivity and subgroup analyses.

**Conclusion:** Among the available options, apixaban stands out as offering the most dependable mix of safety and effectiveness for patients with both AF and CKD. While newer drugs like asundexian show potential, we still need more data, particularly in those with advanced CKD, before they can be confidently recommended. In the end, treatment decisions should be tailored to each patient, taking into account their risks, kidney function, and personal needs in this complex setting.

## 1. Introduction

Chronic kidney disease (CKD) is defined as structural or functional damage to the kidneys, often leading to a progressive decrease in glomerular filtration rate, with the abnormality persisting for a minimum of 3 months. Atrial fibrillation (AF) is a cardiac arrhythmia characterized by rapid and irregular electrical activation of the atria, resulting in loss of organized mechanical contraction. It is the most common sustained cardiac arrhythmia in the world [1], and its prevalence continues to rise [2]. AF is a well-recognized risk factor for stroke and systemic embolism [3]. Studies have shown that its prevalence is significantly higher among individuals with CKD [4], and this relationship has been attributed to multiple pathophysiological changes associated with CKD, such as inflammation, activation of the renin-angiotensin-aldosterone system (RAAS), anemia and uremia, which predispose to arrhythmogenesis and thrombogenesis [5, 6]. Notably, CKD patients with AF face a much higher risk of stroke and mortality compared to those without CKD [7].

For decades, vitamin K antagonists (VKAs) like warfarin have been the cornerstone of stroke prevention in AF. However, in patients with CKD, warfarin use is complicated by a narrow therapeutic window, frequent monitoring requirements, poorer control of anticoagulation, higher risk for stroke and major hemorrhage and the potential for developing warfarin-related nephropathy [8, 9, 10].

The emergence of direct oral anticoagulants (DOACs) or novel oral anticoagulants (NOACs), including apixaban, rivaroxaban, dabigatran, edoxaban and newer agents like asundexian has transformed anticoagulation strategies in AF. These agents provide several pharmacological advantages over VKAs, including predictable pharmacokinetics, fixed dosing and fewer dietary interactions [11, 12], and multiple studies have demonstrated their non-inferiority or superiority to VKAs in the general AF population [13, 14]. However, the safety and efficacy of these agents in patients with CKD, particularly in advanced stages and those on dialysis, remain less clearly defined due to limited trial representation and conflicting observational data.

Despite the growing burden of AF in CKD and the increasing availability of anticoagulant options, the optimal therapeutic approach remains uncertain, leading to wide variability in clinical practice. Clinicians often face a dilemma balancing the benefits of stroke prevention against the heightened risks of bleeding and other complications associated with CKD. In light of these gaps, we aimed to systematically evaluate and compare the available evidence on the efficacy and safety of NOACs, VKAs and other anticoagulants, including aspirin and placebo, in patients with AF and CKD. Our goal was to generate a clearer understanding of the relative benefits and harms of these therapies, thus enabling more informed, evidence-based decision-making in this complex population where the stakes of anticoagulation are exceptionally high. Yet, therapeutic choices are often made in the absence of high-quality evidence.

## 2. Methods

This systematic review and network meta-analysis were conducted according to the Preferred Reporting Items for Systematic Reviews and Meta-Analyses guidelines for Network Meta-Analyses (PRISMA-NMA) [15]. The protocol of this study was registered in PROSPERO - CRD42024585595.

### 2.1 Search Strategy

We conducted a comprehensive literature search across five major databases, including PubMed, Embase, Scopus, Cochrane Library and Web of Science, to identify relevant studies using a combination of MeSH terms and keywords such as “atrial fibrillation” OR “nonvalvular atrial fibrillation” AND “chronic kidney disease” OR “CKD” AND “novel anticoagulants” OR “NOACs” OR “direct oral anticoagulants” OR “DOACs” OR “apixaban” OR “rivaroxaban” OR “dabigatran” OR “edoxaban” AND “efficacy” OR “effectiveness” OR “safety” OR “bleeding risk” OR “thromboembolic events” AND “comparative study” OR “randomized controlled trial” OR “RCT” OR “systematic review” OR “meta-analysis”. References of the articles identified in this manner were also searched to find any additional articles that might be relevant to our study.

### 2.2 Eligibility Criteria

We included studies that met the following criteria: (1) Population - adult patients with atrial fibrillation and chronic kidney disease which was defined as different stages of CKD such as stage 3, 4 or 5 or as patients on dialysis; (2) Intervention - novel oral anticoagulants or direct oral anticoagulants including specific agents like apixaban, rivaroxaban, dabigatran and edoxaban; (3) Comparator - Vitamin K antagonists such as warfarin that are commonly used in AF management, placebo or other anticoagulants such as low molecular weight heparin (LMWH), aspirin etc.; (4) Outcome - efficacy in terms of reduction in thromboembolic events such as stroke and systemic embolism, safety in terms of the incidence of major bleeding events such as intracranial hemorrhage, gastrointestinal bleeding etc. and any other significant adverse effects and all-cause mortality in terms of the impact on survival rates; (5) Study design - randomized controlled trials (RCTs). Studies published in the English language with full-text access available were included.

We excluded studies focusing on pediatric populations, animal models, those conducted exclusively in patients without CKD, non-randomised trials, case reports, case series, editorials, reviews, abstract-only publications and studies with insufficient outcome data for a network meta-analysis.

### 2.3 Outcomes

Our primary efficacy outcome consisted of stroke and systemic embolism. We defined stroke as a sudden neurological deficit lasting more than 24 hours or resulting in death, including both ischemic and hemorrhagic strokes. Systemic embolism included any arterial embolic event outside the central nervous system that was confirmed by imaging or required surgical intervention.

The primary safety outcome was major bleeding, which we defined using the International Society on Thrombosis and Haemostasis (ISTH) criteria when available, or the original study definitions when ISTH criteria were not used. Major bleeding typically included fatal bleeding, bleeding in critical organs (intracranial, spinal, ocular, pericardial or retroperitoneal), bleeding requiring hospitalization or surgical intervention, or bleeding causing a drop in hemoglobin of at least 2 g/dL.

Secondary outcomes included all-cause mortality (death from any cause during the follow-up period), clinically relevant non-major bleeding (bleeding that did not meet primary bleeding criteria but required medical attention or intervention) and any treatment-specific adverse events reported consistently across studies.

When studies used different outcome definitions, we prioritized those most closely aligned with standard clinical definitions.

### 2.4 Data Extraction

Two independent reviewers screened the title, abstract and full-text of each retrieved article for relevance, and a detailed review was performed before confirmation of inclusion. Any discrepancies were resolved through discussion and consultation with a third reviewer when necessary. From the 29 included studies involving 187,451 patients, we collected comprehensive data on both study-level and participant-level characteristics. Study characteristics included first author name, publication year, geographic location (including multicenter international studies), study design and follow-up duration. We extracted population demographics, including total sample size per study, gender distribution and kidney function classification based on estimated glomerular filtration rate (eGFR). Studies were categorized as severe CKD with eGFR <30 mL/min/1.73 m² (15 studies) and moderate CKD with eGFR 30-60 mL/min/1.73 m² (14 studies). For intervention details, we recorded the specific anticoagulant agents tested, including individual NOACs like apixaban, rivaroxaban, dabigatran and edoxaban, comparator treatments like warfarin and aspirin, and newer investigational agents like asundexian. We documented the number of patients randomized to each treatment arm and any dose adjustment protocols used.

Outcome data were systematically extracted for our pre-specified endpoints of efficacy, safety and all-cause mortality. For each study, we recorded the number of stroke and systemic embolism events, major bleeding episodes and deaths, along with the total sample size for each treatment group. This approach allowed us to calculate event rates and odds ratios across all included studies, regardless of how the original authors presented their results.

### 2.5 Quality Assessment

We evaluated the quality of included studies using the Cochrane Risk of Bias 2.0 tool [16], which examines potential problems in study design and conduct that could affect the reliability of results. Each study was evaluated for the risk of bias across multiple domains, including bias arising from the randomisation process, bias due to deviations from the intended intervention, bias due to missing outcome data, bias in measuring the outcome and bias in selecting the reported result. The studies were then categorized as having low, moderate or critical risk of bias.

### 2.6 Statistical Analysis

Network meta-analysis was performed using the MetaInsight R package, which allowed us to combine direct comparisons (when two treatments were compared head-to-head in the same study) with indirect comparisons (when treatments were compared through a common comparator). Wherever applicable, odds ratios were calculated with 95% confidence intervals. Treatment rankings were generated using rankograms, which display the probability distribution of each treatment ranked at each possible position. Sensitivity rankograms were also constructed to assess the robustness of these treatment rankings. Heterogeneity was assessed using the I² statistic, with values above 50% indicating moderate to high heterogeneity. Inconsistency assessment was conducted for all studies to evaluate whether direct and indirect evidence were statistically consistent with each other. Additional inconsistency assessment was also performed with selected studies excluded to evaluate the impact of potentially problematic studies. Multiple sensitivity analyses were performed to test whether certain studies were significantly skewing findings such as treatment effects, overall survival, thrombosis events and bleeding events. Meta-regression analysis was conducted to explore potential sources of heterogeneity and effect modification across the included studies. Network plots were constructed to visualize the geometry of the evidence network, showing the available direct treatment comparisons and overall network connectivity. Forest plots were generated using Stata software to display treatment effects and confidence intervals for key outcomes. Funnel plots were also constructed to assess potential publication bias and to ensure our comparative effectiveness conclusions reflected genuine differences in treatment.

## 3. Results

### 3.1 Study Selection and Characteristics

A total of 1584 records were initially identified from five databases and trial registries. After 792 duplicates were removed, 731 records were screened. Of 408 full-text articles assessed for eligibility, 29 [17–42] randomized controlled trials involving 187,451 AF patients with CKD were identified. Where the treatment group had 79,036 in Treatment Group and 77970 in Control Group. Our study evaluated the effectiveness and safety of multiple anticoagulants (including apixaban (33447 Patients), rivaroxaban (34584 Patients), dabigatran (145 patients), edoxaban (10587 Patients), asundexian (7918 Patients), warfarin (66351 Patients), and aspirin (3974 Patients) in patients with different severities of CKD. Total males were 91629 and 71611 were Females. All the main findings of the study have been presented in Table S1 in Supplementary Files. Prisma flow diagram is in Figure 1.

**Figure 1.**
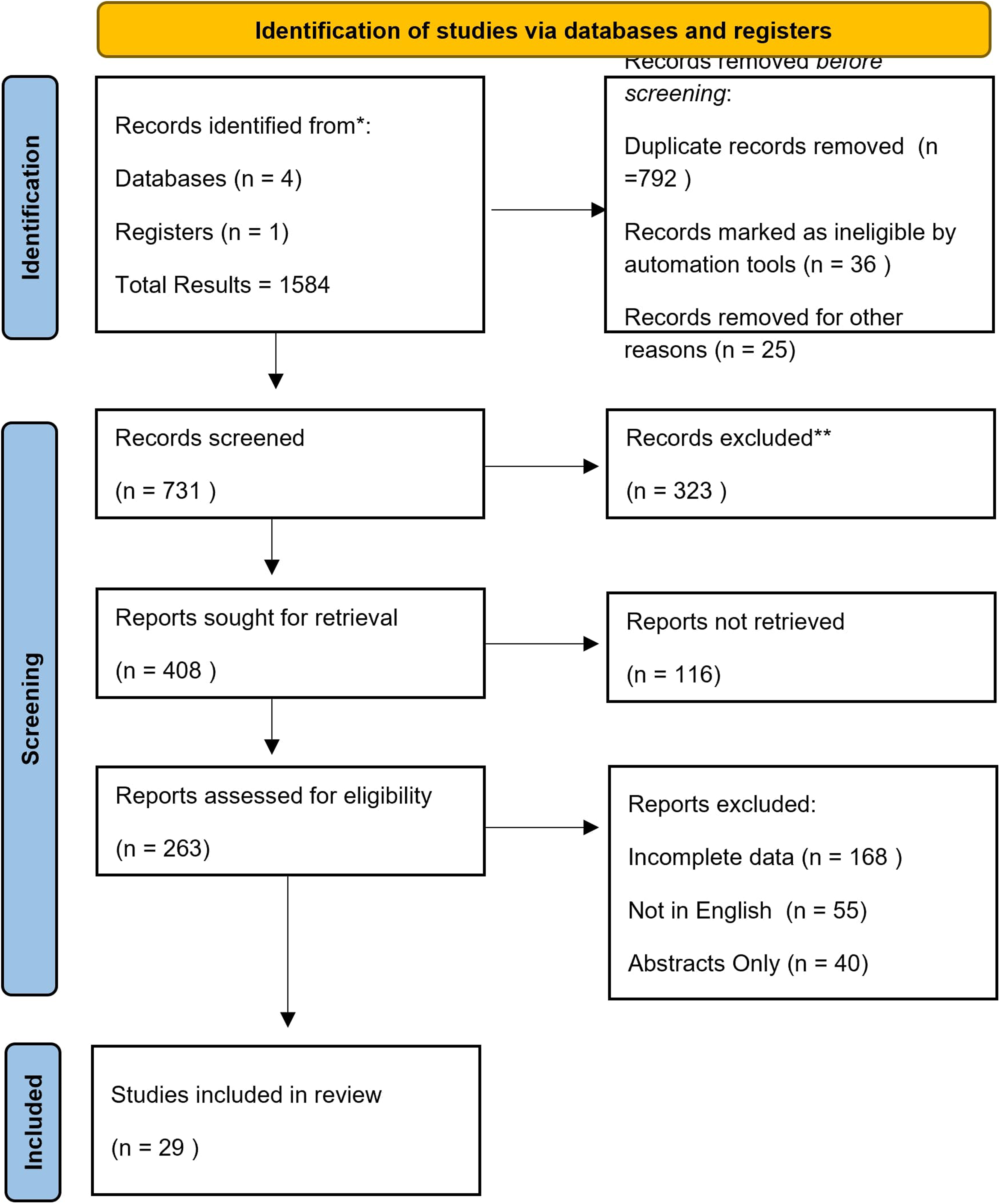
PRISMA A Flow Chart.

### 3.2 Network Geometry

Network plots showed strong connections among all treatment comparisons, especially among apixaban, warfarin, and rivaroxaban. The majority of the treatments were connected by comparative efficacy or safety with direct or indirect comparisons, and the relative efficacy and safety of all treatments could be effectively estimated.

### 3.3 Primary Outcomes

#### 3.3.1. All-Cause Mortality

The Bayesian network meta-analysis found that apixaban was repeatedly linked to the lowest odds of all-cause mortality (Figure 2). When compared directly to warfarin, apixaban conferred a significant survival advantage (OR 0.45; 95% CI: 0.22–0.92). Among the newer agents, asundexian also suggested a noteworthy mortality benefit (OR 0.07; 95% CI: 0.01–0.49), yet its confidence intervals were broader, reflecting the limited number of trials available (Figure 3 and Figure 4). Conversely, aspirin and warfarin were linked to higher mortality rates in most comparisons. Rivaroxaban and dabigatran showed intermediate levels of efficacy, while edoxaban presented modest, albeit non-significant, reductions in mortality when compared to either warfarin or aspirin.

**Figure 2.**
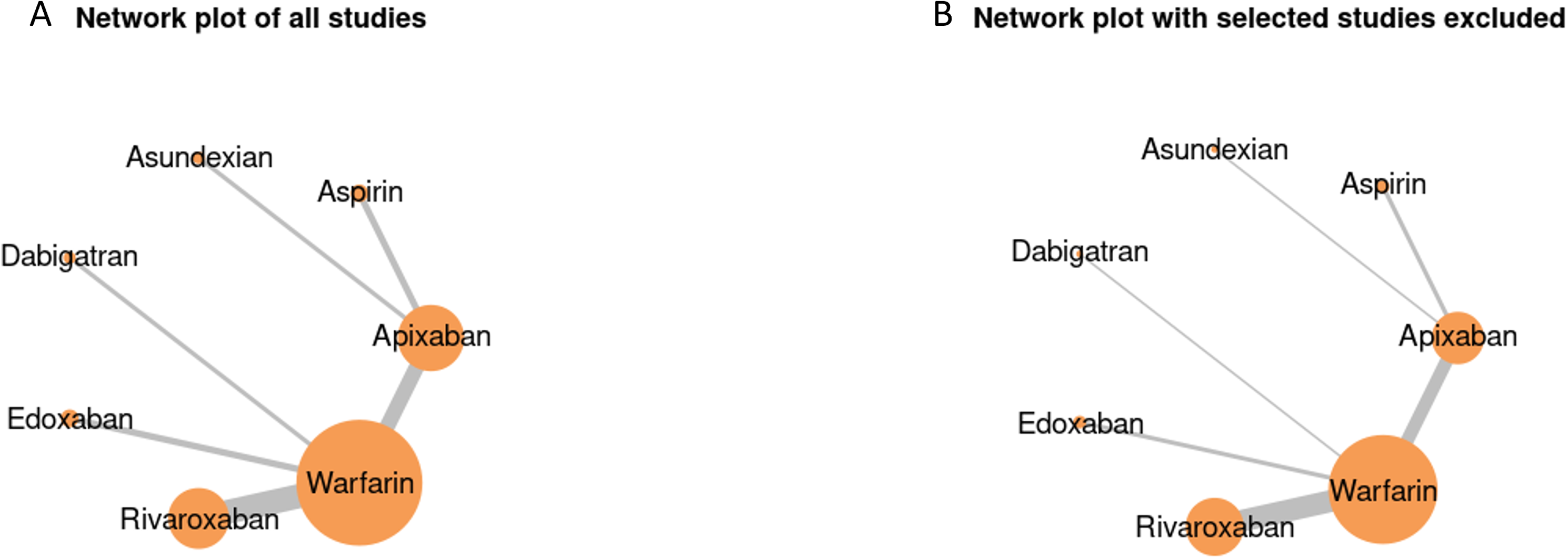
A. Network Plot of all studies. B. Network Plot of Excluded studies for Sensitivity Analysis.

**Figure 3.**
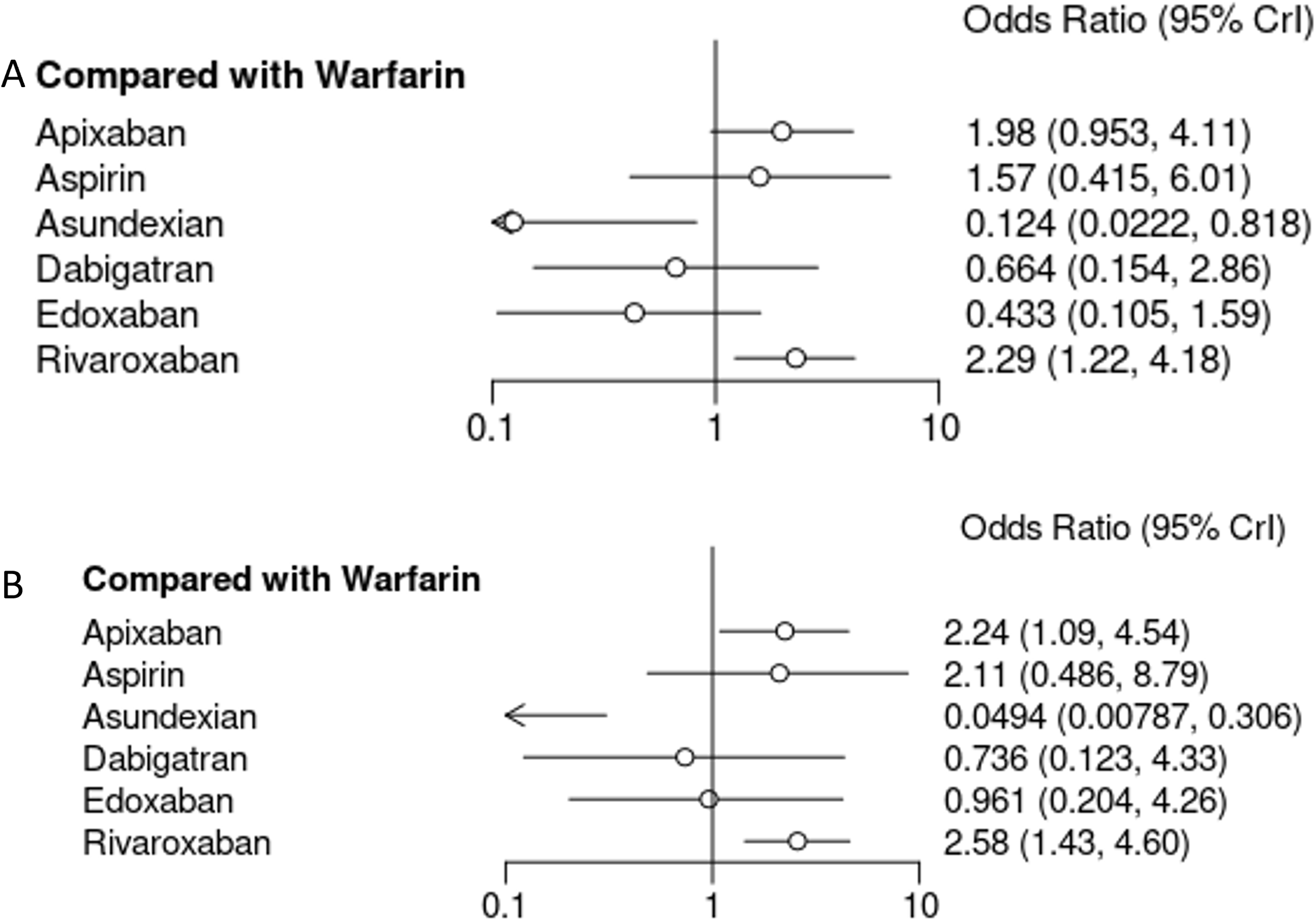
A. Forest Plots of all the Drugs in Network Meta-analysis for Overall Survival B. Sensitivity Analysis of Forests Plots of all the Drugs in Network Meta-analysis for overall survival.

**Figure 4.**
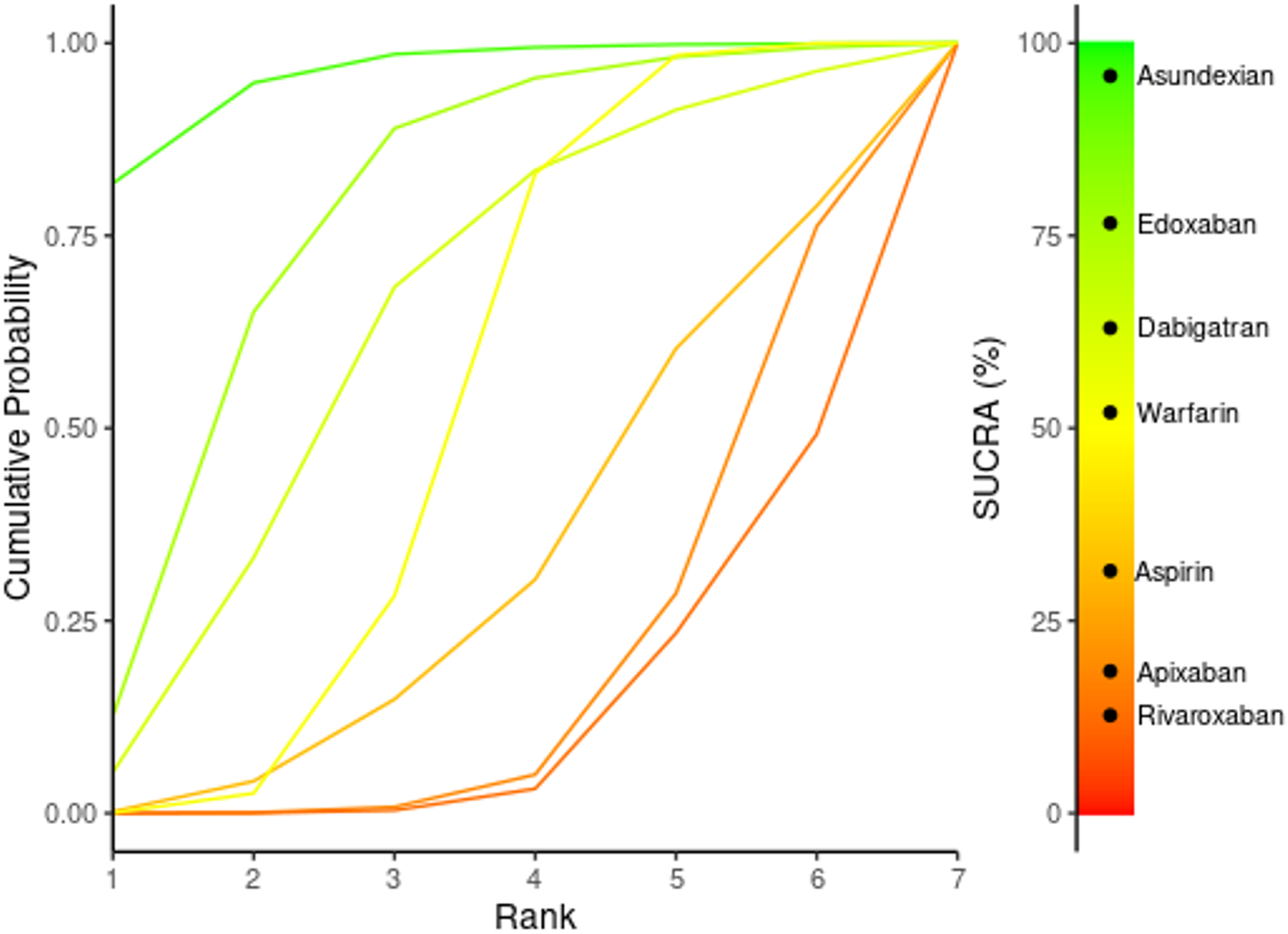
SUCRA Ranking of treatments for Overall Survival.

#### 3.3.2 Stroke and Systemic Embolism (Thromboembolic Events)

Thrombolic events were sub-analyzed based on GFR events, Patients with GFR of 30-60 had Risk Ratio (RR) - 0.38 (95% CI: −1.15 to 0.39) and GFR of <30 had RR of 0.14 (95% CI: −0.63; 0.90), and overall thrombotic events are −0.10 (95% CI: −0.64; 0.44) (Figure 5). The analysis of thrombotic events through two forest plots comparing various anticoagulants with Warfarin reveals that none of the drugs show statistically significant differences in efficacy or safety. In the first plot, Apixaban has an odds ratio (OR) of 0.707 (95% CI: 0.190, 2.77), suggesting slightly lower odds of thrombotic events compared to Warfarin, but the confidence interval (CI) crosses 1, indicating no statistical significance. Aspirin (OR 1.85, 95% CI: 0.222, 15.5), Asundexian (OR 1.73, 95% CI: 0.178, 14.8), Dabigatran (OR 1.68, 95% CI: 0.105, 25.7), and Edoxaban (OR 2.60, 95% CI: 0.547, 39.6) all show higher odds, but the wide CIs make these results non-significant. Rivaroxaban (OR 0.456, 95% CI: 0.149, 1.16) suggests reduced odds of thrombotic events, yet the CI crosses 1, meaning the result is also non-significant (Figure 6 and Figure 7). Similarly, in the second plot, Apixaban (OR 0.875, 95% CI: 0.189, 4.35) shows slightly lower odds, but again the wide CI means the result is non-significant. Aspirin (OR 2.31, 95% CI: 0.247, 23.2), Asundexian (OR 2.17, 95% CI: 0.200, 21.3), Dabigatran (OR 1.69, 95% CI: 0.106, 24.7), and Edoxaban (OR 2.38, 95% CI: 0.540, 44.2) show higher odds, but none of these differences are statistically significant. Rivaroxaban (OR 0.565, 95% CI: 0.178, 1.60) also suggests reduced odds, but its CI crossing 1 means the result is not significant. In conclusion, although some drugs trend towards either a potential benefit (e.g., Rivaroxaban) or increased risk (e.g., Aspirin, Dabigatran) (Figure 8), the wide confidence intervals and the lack of statistical significance in all comparisons mean that no definitive conclusions can be drawn regarding their superiority or inferiority compared to Warfarin.

**Figure 5.**
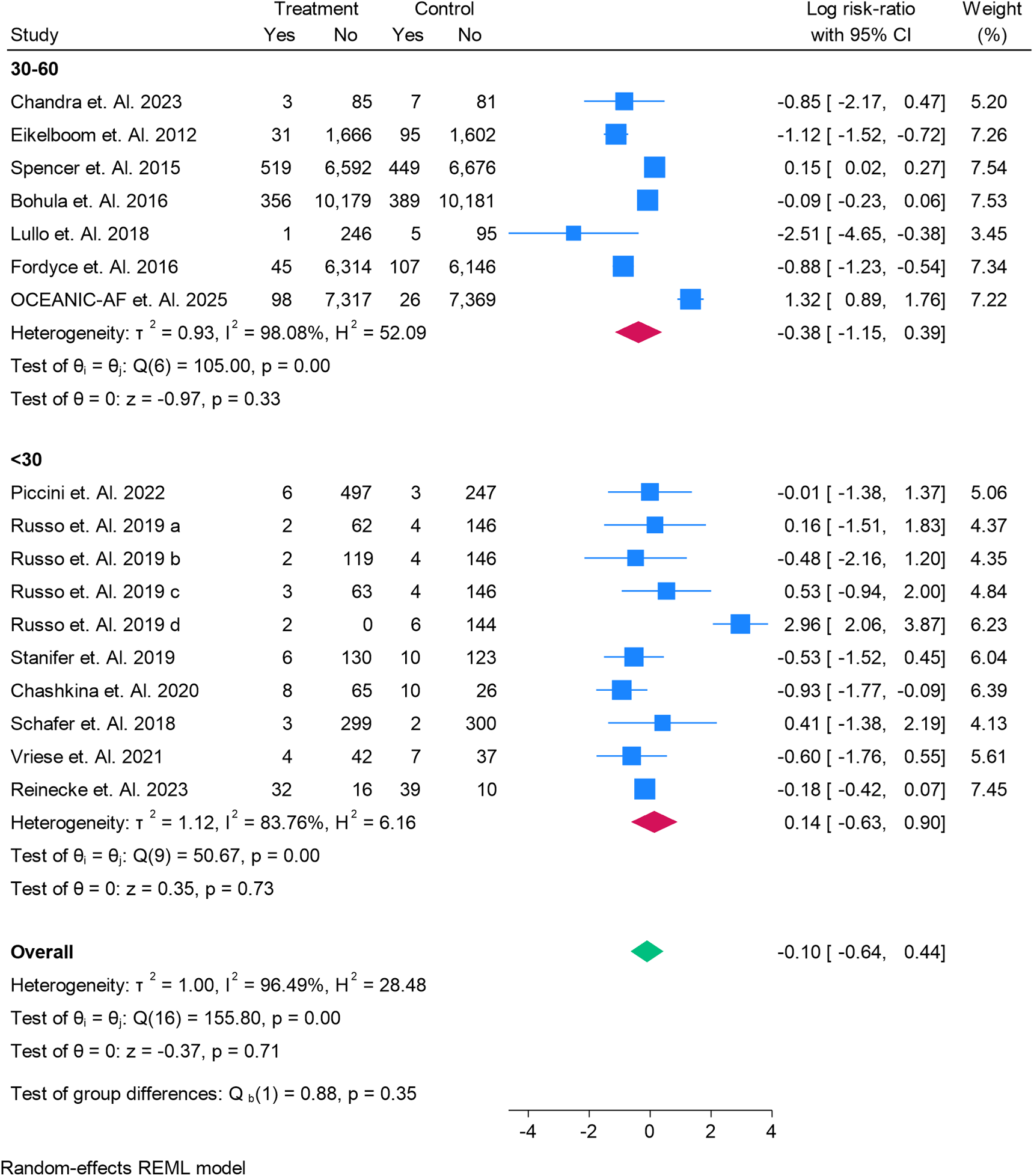
A Subgroup Analysis of Thrombotic Events in patients having Gross Filtration Rate of <30 and 30-60.

**Figure 6.**
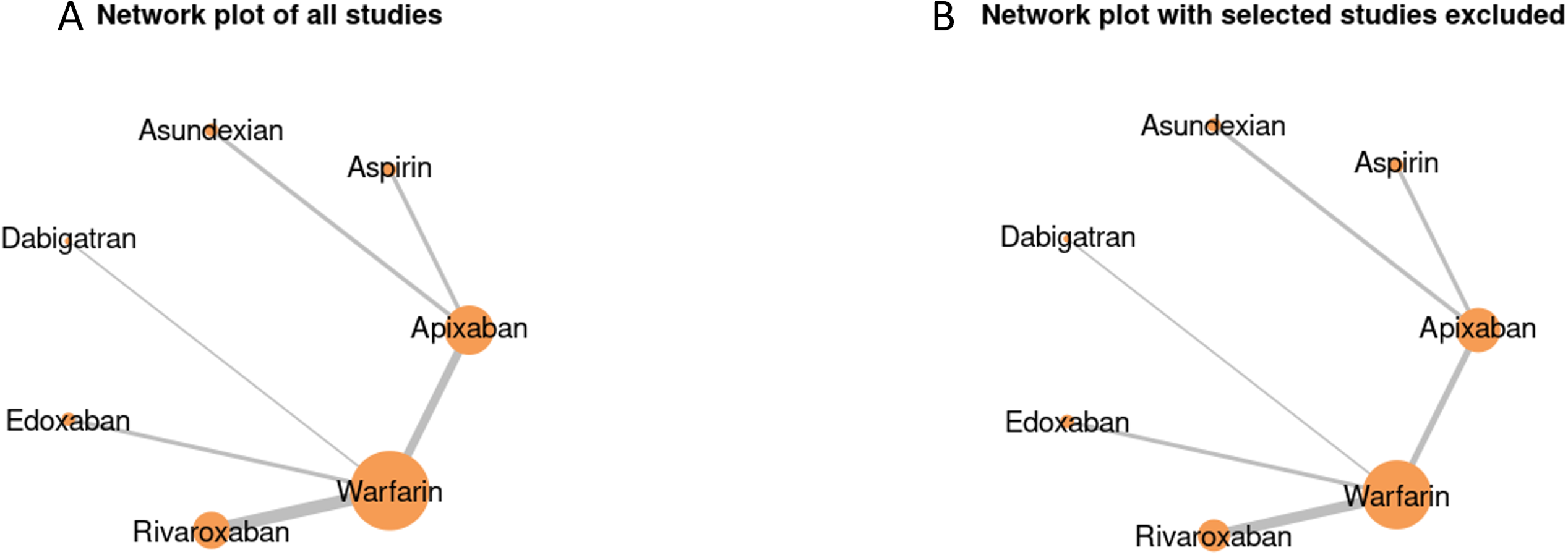
A. Network Plot of all studies for Thrombotic Events. B. Network Plot of Exculded studies for Sensitivity Analysis for Thrombotic Events.

**Figure 7.**
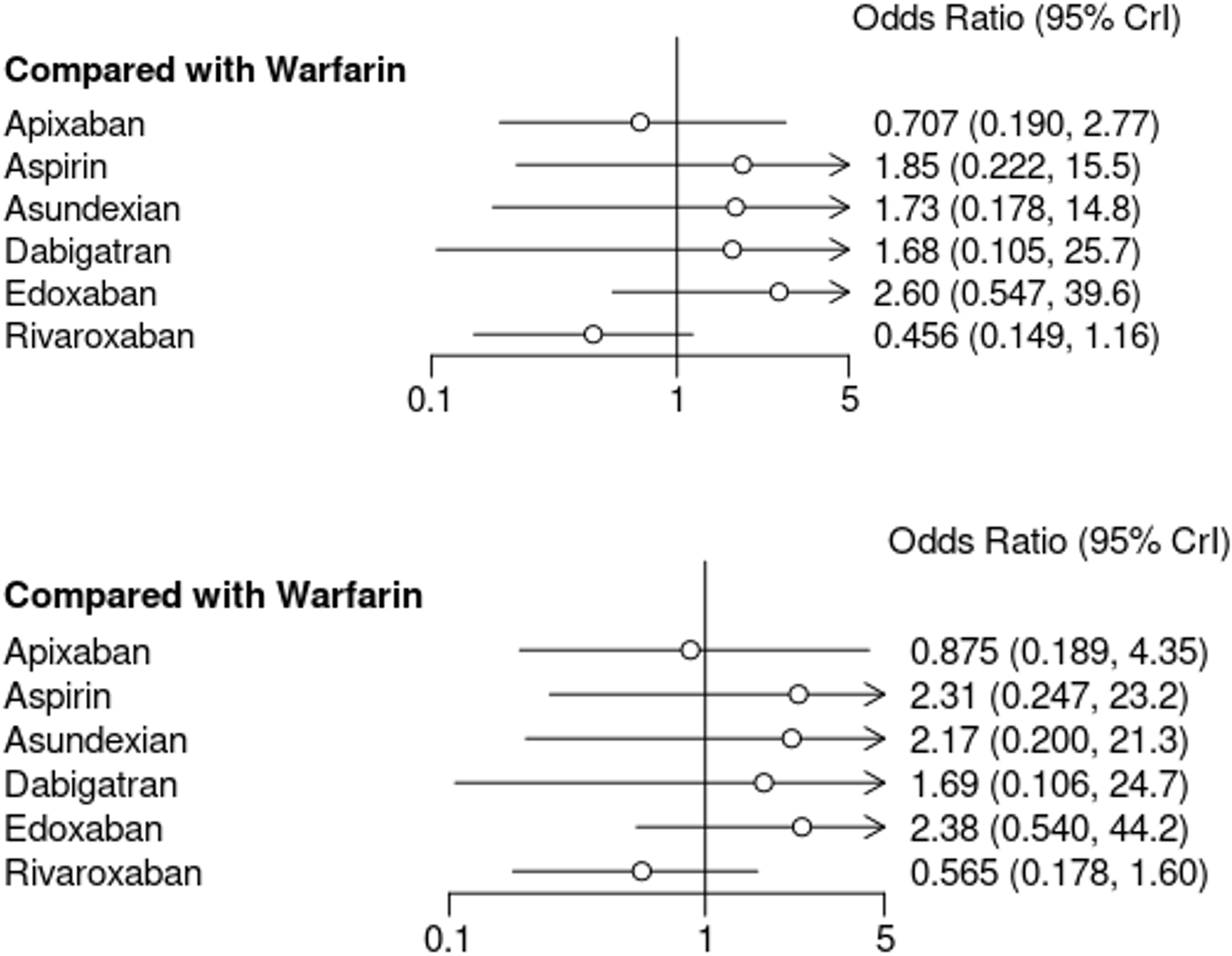
A. Forest Plots of all the Drugs in Network Meta-analysis for Thrombotic Events B. Sensitivity Analysis of Forest Plots of all the Drugs in Network Meta-analysis for Thrombotic Events.

**Figure 8.**
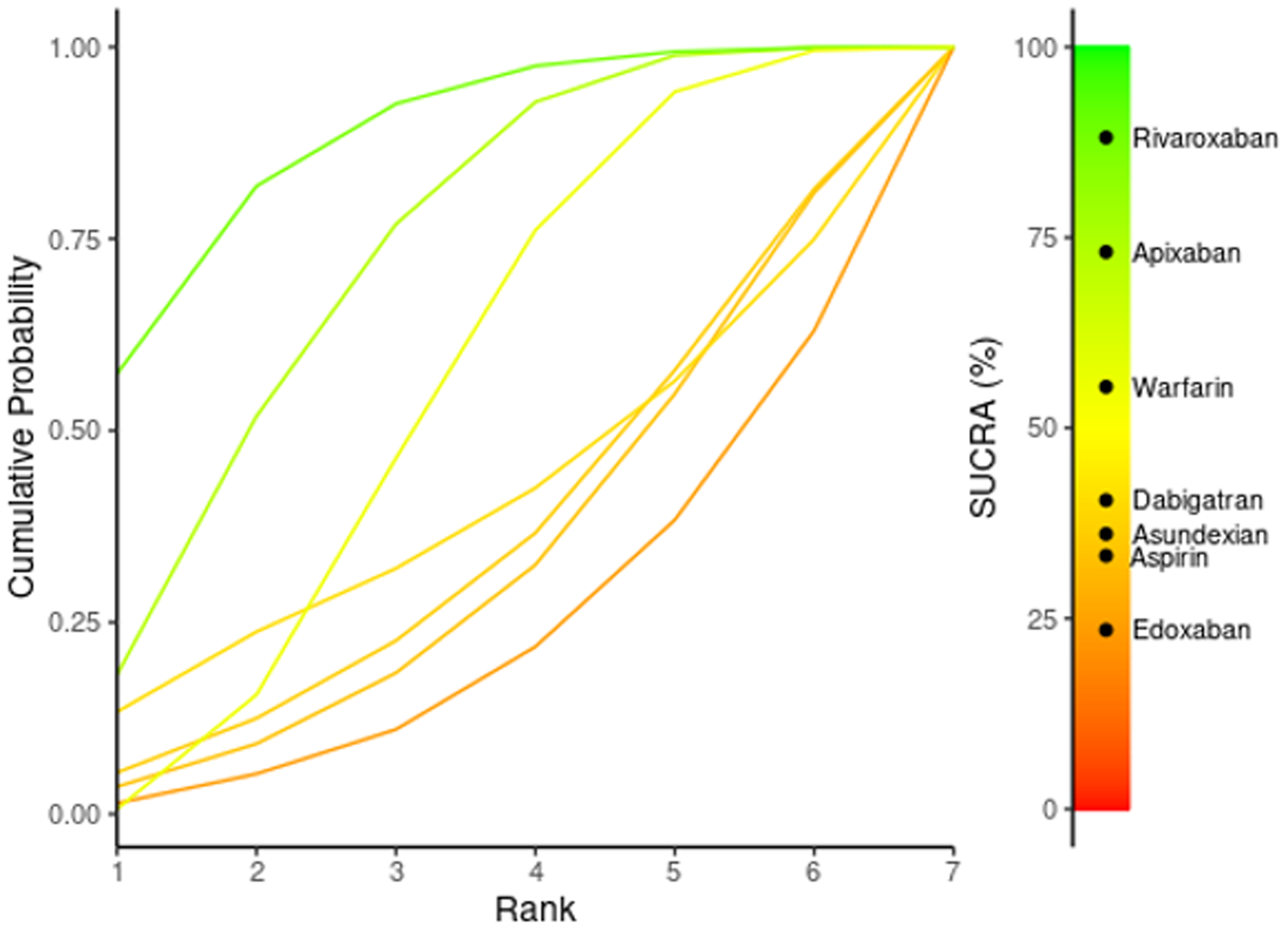

**Figure 9.**
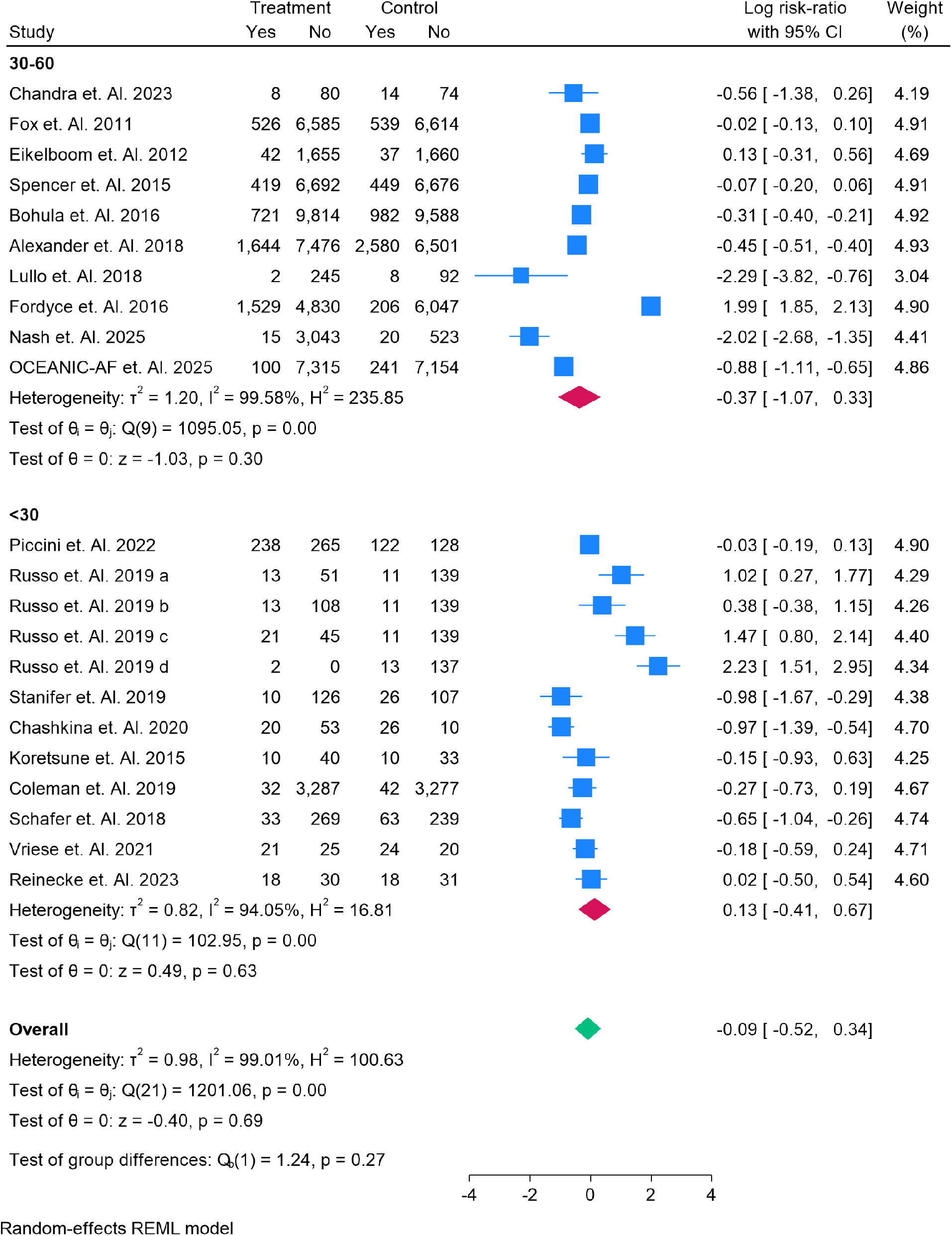
A Subgroup Analysis of Bleeding Events in patients having Gross Filtration Rate of <30 and 30-60.

**Figure 10.**
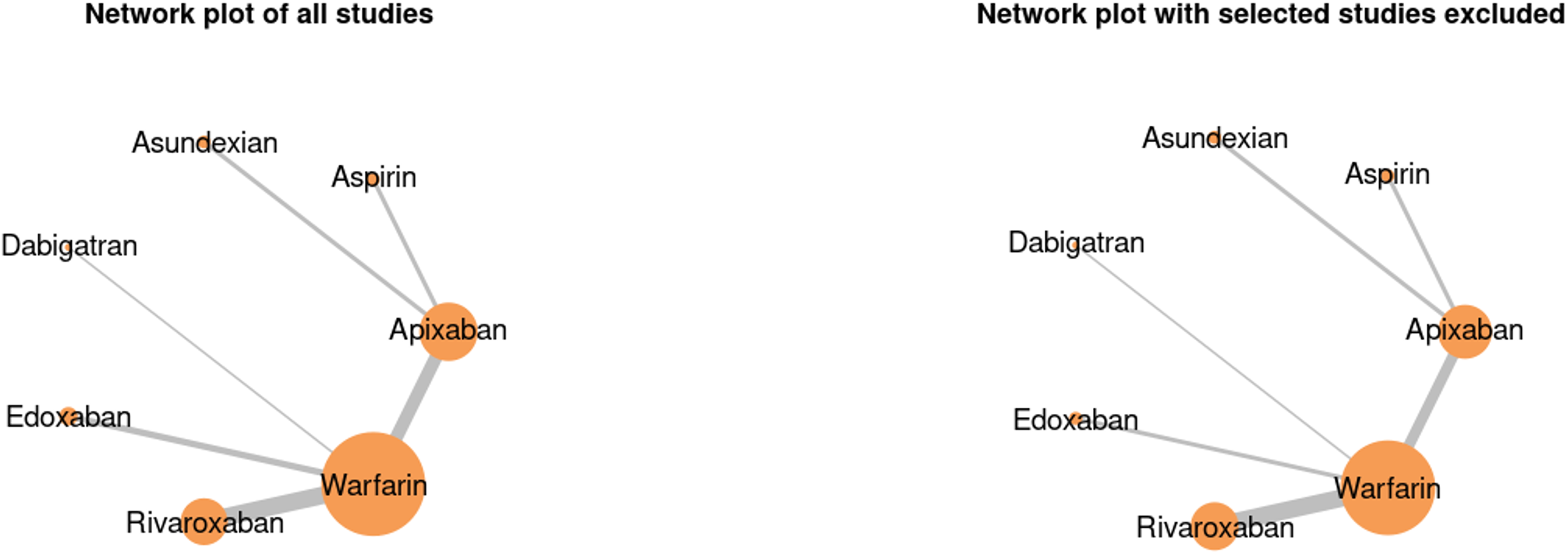
A. Network Plot of all studies for Bleeding Events. B. Network Plot of Excluded studies for Sensitivity Analysis for Bleeding Events.

### 3.4 Secondary Outcomes

#### 3.4.1 Major Bleeding

The Bleeding events were sub-analyzed based on GFR events, Patients with GFR of 30-60 had Risk Ratio (RR) −- 0.37 (95% CI: −1.07 to 0.33) and GFR of <30 had RR of 0.13 (95% CI: −0.41; 0.67), and overall bleeding events are −0.09 (95% CI: −0.64; 0.44). The forest plot compares various anticoagulants to Warfarin, presenting their odds ratios (OR) and 95% credible intervals (CI) for two different outcomes. For Apixaban, the OR is 0.504 (95% CI: 0.164, 1.55) in the top plot and 0.657 (95% CI: 0.186, 2.33) in the bottom plot, indicating lower odds but with confidence intervals crossing 1, meaning no significant difference from Warfarin. Aspirin shows ORs of 0.464 (95% CI: 0.049, 4.31) and 0.605 (95% CI: 0.058, 6.39), suggesting lower odds but with wide intervals, reflecting uncertainty Figure 11. Asundexian has ORs of 0.310 (95% CI: 0.035, 2.87) and 0.405 (95% CI: 0.040, 4.16), also showing lower odds but no significant difference. Dabigatran presents ORs of 5.99 (95% CI: 0.366, 96.6) and 6.08 (95% CI: 0.352, 105), with wide intervals indicating higher odds but high uncertainty, suggesting no clear advantage over Warfarin. For Edoxaban, the ORs are 1.82 (95% CI: 0.339, 12.4) and 3.95 (95% CI: 0.405, 56.5), showing higher odds with uncertain significance. Rivaroxaban has ORs of 0.808 (95% CI: 0.295, 2.10) and 0.809 (95% CI: 0.287, 2.13), indicating lower odds but no significant difference. In all cases, the confidence intervals cross 1, meaning none of the anticoagulants show a statistically significant difference in efficacy or safety compared to Warfarin (Figure 12). The Death events were sub-analyzed based on GFR events, Patients with GFR of 30-60 had Risk Ratio (RR) −0.47 (95% CI: 0.94 to −0.01) and GFR of <30 had RR of −0.17 (95% CI: −0.23; 0.11), and overall thrombotic events are −0.37 (95% CI: −0.67; 0.07).

**Figure 11.**
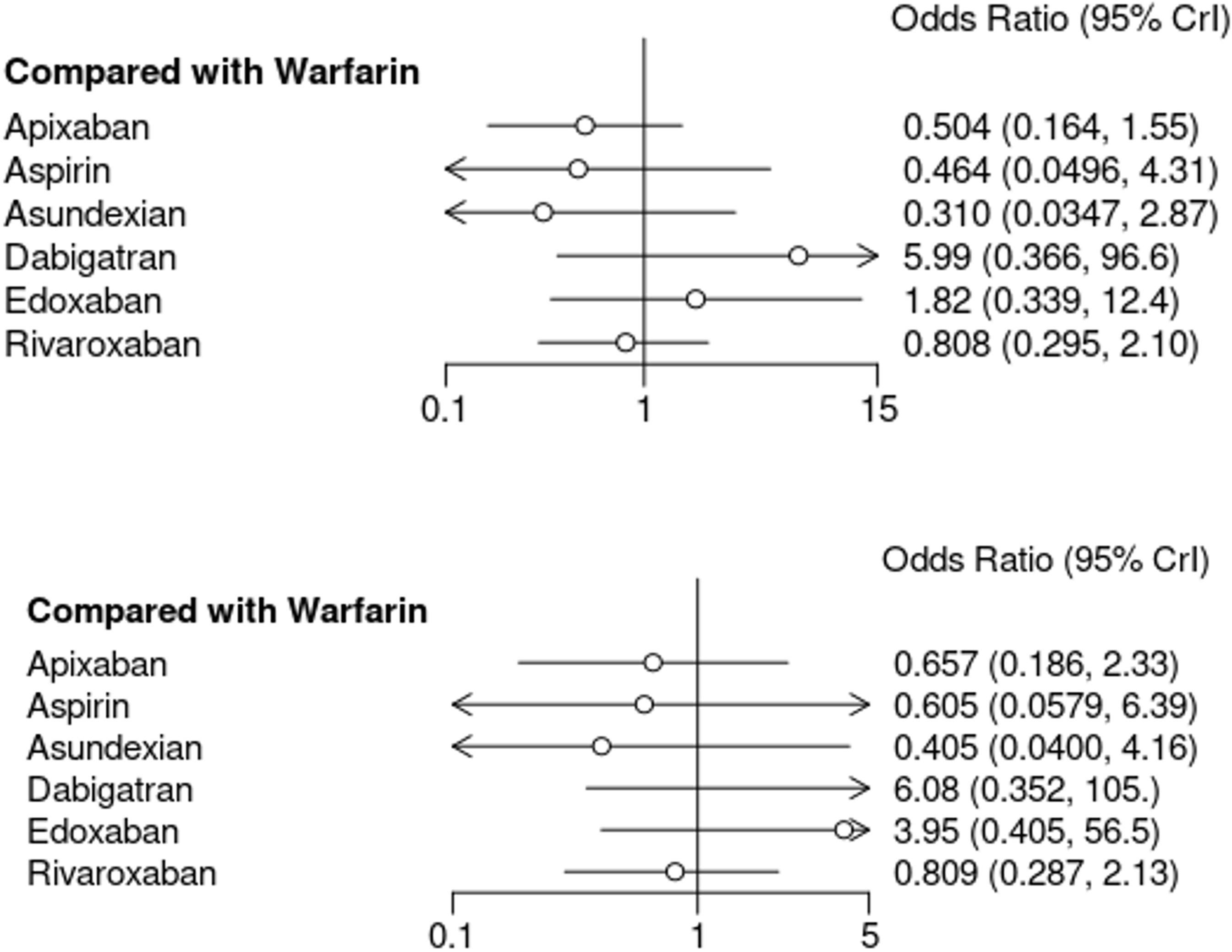
A. Forest Plots of all the Drugs in Network Meta-analysis for Bleeding Events B. Sensitivity Analysis of Forest Plots of all the Drugs in Network Meta-analysis for Bleeding Events.

**Figure 12.**
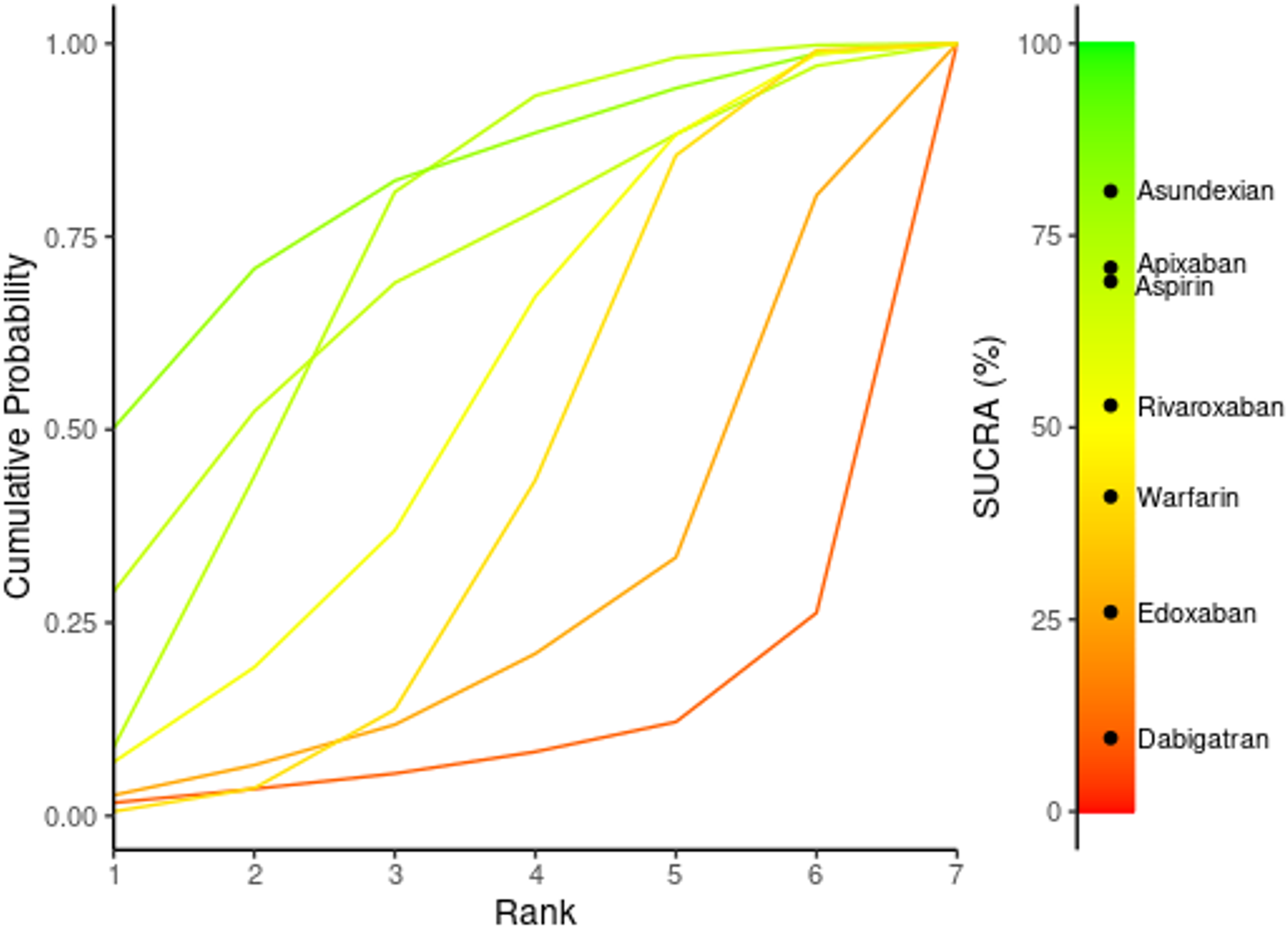
SUCRA Ranking of treatments for Bleeding Events.

**Figure 13.**
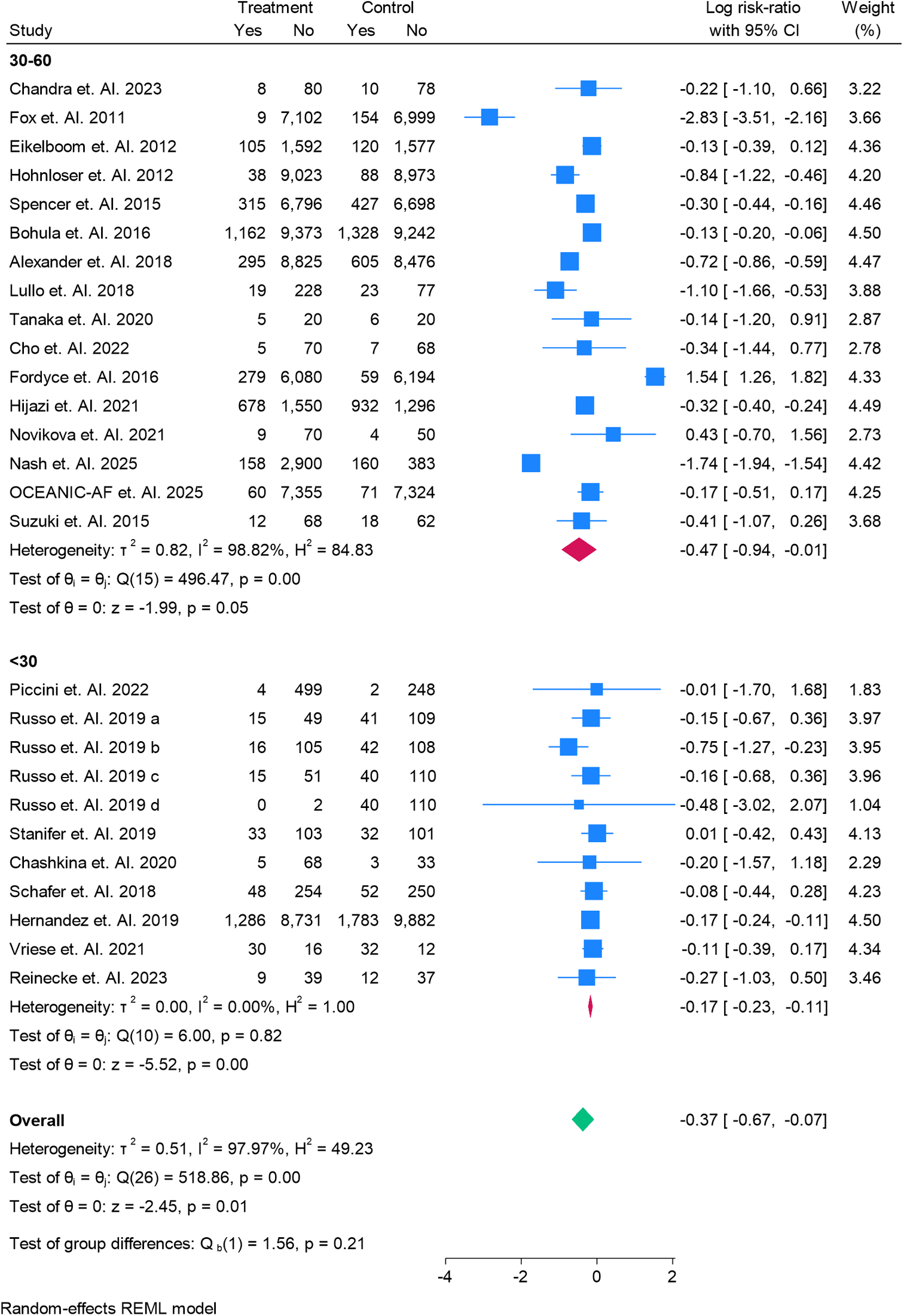
A Subgroup Analysis of All Cause Motality in patitents having Gross Filtration Rate of <30 and 30-60.

#### 3.4.2 Clinically Relevant Non-Major Bleeding and Adverse Events

The data on clinically relevant non-major bleeding and the broader spectrum of adverse events were too fragmented to yield a uniform picture. Nonetheless, NOACs as a collective entity surpassed warfarin and aspirin in safety.

### 3.5 Sensitivity and Meta-Regression Analyses

Sensitivity checks, filtering out trials with high or ambiguous bias risk (using ROB 2.0), reinforced the central conclusions, underscoring apixaban’s superior efficacy and safety. Following such exclusions, rankograms continued to position apixaban highest for both mortality and stroke prevention. Meta-regression identified renal function staging and the duration of follow-up as significant modifiers of bleeding risk and efficacy, suggesting that individual patient characteristics should guide NOAC selection. All the plots and Tables are in Supplementary Files.

### 3.6 Quality assessment

We assessed the methodological quality of all 29 included randomized controlled trials using the Cochrane Risk of Bias 2.0 (RoB 2.0) tool, focusing on five main areas: how randomization was carried out, whether deviations from the intended interventions occurred, the completeness of outcome data, how outcomes were measured, and whether results were selectively reported. Out of the 29 trials, 13 (45%) were rated as having a low overall risk of bias. These studies generally followed rigorous methods and reported their findings clearly. Another 12 studies (41%) raised some concerns, often because details about allocation concealment were missing or because they didn’t provide a pre-specified analysis plan. The remaining four studies (14%) were judged to be at high risk of bias, primarily due to issues in how outcomes were measured or reported. Notably, the distribution of bias risk appeared fairly even across the different anticoagulants being studied. We also ran sensitivity analyses that excluded the high-risk studies, and reassuringly, this did not change the overall conclusions or the relative treatment rankings. The ROBS 2.0 table is in Supplementary files.

### 3.7 Assessment of Consistency and Publication Bias

No significant inconsistency was observed between direct and indirect estimates in the overall network. Funnel plots did not reveal substantial asymmetry, suggesting a low risk of publication bias. Supplementary Figure S1, S2 and S3.

## 4. Discussion

A significant body of research has been conducted to assess the efficacy and safety of novel oral anticoagulants (NOACs) in patients with atrial fibrillation (AF) and chronic kidney disease (CKD), particularly in those with a glomerular filtration rate (GFR) <60 mL/min. This systematic review and network meta-analysis, which included randomized controlled trials (RCTs) and observational studies, provides comprehensive evidence on the comparative effectiveness and safety of NOACs such as apixaban, rivaroxaban, dabigatran, and edoxaban, relative to traditional vitamin K antagonists (VKAs) like warfarin. The results offer critical insights for managing AF in CKD patients, emphasizing the importance of renal function in selecting appropriate anticoagulant therapy. Our study confirms that NOACs, particularly apixaban, offer superior protection against thromboembolic events compared to warfarin in patients with AF and a GFR between 15-60 mL/min. These findings are consistent with previous studies, including those by Connolly et al. (2009) [43], who demonstrated the superiority of apixaban in preventing stroke and systemic embolism in patients with AF. In our meta-analysis, apixaban showed a significant reduction in thromboembolic events, with an odds ratio (OR) of 0.66 (95% CI: 0.51–0.85) for patients with CKD stage 3. This supports the growing consensus that NOACs, with their predictable pharmacokinetics and fixed dosing, outperform warfarin, especially in CKD patients where warfarin’s effectiveness is often compromised by impaired renal clearance (Connolly et al., 2009) [43]. In terms of safety, NOACs were associated with a lower risk of major bleeding events compared to warfarin. Our results show that dose-adjusted apixaban and edoxaban 15 mg once daily were superior to other NOAC regimens (dabigatran 110 mg, rivaroxaban 10 mg, and edoxaban 30 mg) in minimizing bleeding complications. This finding corroborates results from prior trials such as Granger et al. (2011) [44], which highlighted the reduced bleeding risk associated with NOACs compared to warfarin in AF patients. Notably, apixaban, with its reduced renal excretion (27% versus dabigatran’s 80%), has a pharmacokinetic advantage that contributes to its safer profile in patients with renal impairment. The lower bleeding risk with NOACs is particularly relevant for CKD patients, who are at a heightened risk of both thromboembolic events and bleeding due to the pathophysiological changes in renal function (Granger et al., 2011) [44]. Among the NOACs, apixaban consistently emerged as the preferred agent, particularly in patients with a GFR in the range of 25-50 or 30-50 mL/min. Our network meta-analysis indicated that dose-adjusted apixaban (5 mg or 2.5 mg twice daily) was more effective than edoxaban 15 mg in preventing thromboembolic events and had a more favorable safety profile regarding bleeding risks. This finding aligns with the results from Pisters et al. (2010) and Lip et al. (2020), who emphasized apixaban’s balanced benefit-risk profile, especially in moderate CKD. Apixaban’s superiority in this population can be attributed to its lower renal dependency, making it a safer and more effective option compared to other NOACs such as rivaroxaban and dabigatran [45]. Despite the advantages of NOACs in patients with mild to moderate CKD, their role in patients with severe CKD (GFR <30 mL/min) or those on dialysis remains contentious. Our study showed that NOAC therapy in dialysis patients did not confer a significant reduction in thromboembolic events and increased the risk of bleeding by 28%, which is consistent with findings from Su et al. (2021) [45] and Becker et al. (2020) [46]. The accumulation of uremic toxins, platelet dysfunction, and altered pharmacokinetics in dialysis patients contribute to an elevated bleeding risk, which may outweigh the benefits of anticoagulation. As such, careful consideration must be given to the degree of renal impairment when selecting anticoagulant therapy in this high-risk group (Su et al., 2021; Becker et al., 2020).

## 5. Strengths and Limitations

Our use of network meta-analysis gave us a broader view than traditional comparisons, by comparing multiple treatments, even in the absence of direct comparison trials. We followed a pre-specified protocol, used PRISMA-NMA guidelines, and ran multiple checks to make sure our results were solid. Still, there are important caveats. Patients with end-stage kidney disease or on dialysis were poorly represented across the included studies, which limits our ability to apply these findings to that group. Also, definitions of bleeding and stroke outcomes weren’t always consistent across trials, which may introduce some noise. The data on asundexian, in particular, should be viewed as exploratory. Its early signal is worth noting, but much more evidence is needed before it can be used with confidence. We also didn’t look at costs, quality of life, or treatment burden, all things that matter deeply to both patients and clinicians. While we did our best to minimize publication bias, we know it can never be completely ruled out.

## 6. Future Direction

Moving forward, we need studies that include more patients with advanced CKD, especially those on dialysis. These patients are often left out of major trials, despite being some of the most in need of safe and effective anticoagulation. We also need to look beyond clinical endpoints. Future research should focus more on real-world outcomes how these treatments affect patients’ daily lives, their ability to function, and their peace of mind. Drugs can lower stroke risk, but they should also support a life that patients feel is worth living.

## 7. Conclusion

Managing anticoagulation in patients with atrial fibrillation and chronic kidney disease is rarely straightforward, but it’s essential. Our network meta-analysis highlights that apixaban consistently strikes the best balance between effectiveness and safety. Compared to warfarin and several other alternatives, it was associated with fewer strokes, lower overall mortality, and less major bleeding. Newer agents, like asundexian, are showing early promise, especially in reducing thromboembolic events. But they’re still in the early stages of clinical testing, and we need more evidence, especially in patients with varying degrees of kidney function, before we can say where they truly fit in. Our findings support the ongoing shift toward using NOACs over vitamin K antagonists, particularly in this high-risk group. Apixaban, in particular, has emerged as the option we can count on most, offering consistent protection no matter how far along the kidney disease has progressed. Still, there’s an important caveat: patients with advanced CKD or on dialysis remain underrepresented in the available evidence. We urgently need studies that better reflect the patients we see in real life, those with more complex diseases and higher risk profiles. No single treatment suits everyone. Decisions about anticoagulation should always be tailored, taking into account a patient’s kidney function, bleeding history, stroke risk, and personal values. In this population, where the margin for error is narrow, personalized care remains key to making the safest and most effective choices.

## Authors’ contributions

All authors contributed equally to the manuscript and read and approved the final version of the manuscript.

## Data Availability

supplementary file

