## supplementary file for "Comparative Efficacy and Safety of Novel Oral Anticoagulants in Atrial Fibrillation with Chronic Kidney Disease: A Systematic Review, Meta-Regression, and Network Meta-Analysis"

Supplementary Figure

Search Strategy

(“atrial fibrillation” OR “nonvalvular atrial fibrillation”) AND (“chronic kidney disease” OR “CKD”) AND (“novel anticoagulants” OR “direct oral anticoagulants” OR “DOACs” OR “apixaban” OR “rivaroxaban” OR “dabigatran” OR “edoxaban”) AND (“efficacy” OR “effectiveness” OR “safety” OR “bleeding risk” OR “thromboembolic events”)

Table S1. Demographics and Summary Table

| **Ref. Number** | **Author Year** | **Country** | **Total** | **Male** | **Female** | **Treatment Group** | **Patients in Treatment Group** | **Control Group** | **Patients in Control Group** | **GRADE** | **Main finding** |
| --- | --- | --- | --- | --- | --- | --- | --- | --- | --- | --- | --- |
| 17 | Piccini et. Al. 2022 | Multicenter | 862 | 446 | 309 | Asundexian | 503 | Apixaban | 250 | High | he PACIFIC-AF trial found that asundexian (20 mg and 50 mg) resulted in significantly fewer bleeding events compared to apixaban, with similar thrombotic event rates. |
| 18 | Russo et. Al. 2019 a | Italy | 1053 | 329 | 724 | Apixaban | 64 | Warfarin | 150 | High | The study found that direct oral anticoagulants (DOACs) were associated with a lower incidence of all-cause mortality compared to vitamin K antagonists (VKAs) in octogenarians with atrial fibrillation, although the rates of thromboembolic and bleeding events were similar between the two groups |
| 19 | Russo et. Al. 2019 b | Italy | 1053 | 329 | 724 | Rivaroxaban | 121 | Warfarin | 150 | Moderate | The main finding of the study is that apixaban resulted in fewer major bleeding events compared to warfarin in patients with atrial fibrillation and advanced chronic kidney disease (CrCl 25–30 mL/min). |
| 20 | Russo et. Al. 2019 c | Italy | 1053 | 329 | 724 | Dabigatran | 66 | Warfarin | 150 | High | The main finding of the study is that apixaban resulted in fewer major bleeding events compared to warfarin in patients with atrial fibrillation and advanced chronic kidney disease (CrCl 25–30 mL/min). |
| 21 | Russo et. Al. 2019 d | Italy | 1053 | 329 | 724 | Edoxaban | 2 | Warfarin | 150 | Moderate | The main finding of the study is that apixaban resulted in fewer major bleeding events compared to warfarin in patients with atrial fibrillation and advanced chronic kidney disease (CrCl 25–30 mL/min). |
| 22 | Stanifer et. Al. 2019 | Multicenter | 269 | 106 | 163 | Apixaban | 136 | Warfarin | 133 | High | The main finding of the study is that apixaban resulted in fewer major bleeding events compared to warfarin in patients with atrial fibrillation and advanced chronic kidney disease (CrCl 25–30 mL/min). |
| 23 | Chandra et. Al. 2023 | India | 192 | 89 | 87 | Apixaban | 88 | Warfarin | 88 | Moderate | The main finding of the study is that **apixaban** therapy resulted in **lower incidences of major bleeding, stroke, and thromboembolism** compared to **warfarin** in patients with atrial fibrillation and chronic kidney disease (stage 3-5) |
| 24 | Chashkina et. Al. 2020 | Russia | 109 | 60 | 49 | Rivaroxaban | 73 | Warfarin | 36 | High | The main finding of the study is that apixaban resulted in fewer major bleeding events compared to warfarin in patients with atrial fibrillation and advanced chronic kidney disease (CrCl 25–30 mL/min). |
| 25 | Fox et. Al. 2011 | Multicenter | 14264 | 8803 | 5461 | Rivaroxaban | 7111 | Warfarin | 7153 | High | The main finding of the study is that apixaban resulted in fewer major bleeding events compared to warfarin in patients with atrial fibrillation and advanced chronic kidney disease (CrCl 25–30 mL/min). |
| 26 | Eikelboom et. Al. 2012 | Multicenter | 5595 | 3301 | 2294 | Apixaban | 1697 | Aspirin | 1697 | High | The main finding of the trial is that **apixaban significantly reduced the risk of stroke and systemic embolism compared to aspirin in patients with stage III chronic kidney disease, without an increased risk of major bleeding.** |
| 27 | Hohnloser et. Al. 2012 | Multicenter | 18122 | 9061 | 9061 | Apixaban | 9061 | Warfarin | 9061 | High | The main finding of the study is that apixaban reduces the rates of stroke, death, and major bleeding compared to warfarin in patients with atrial fibrillation, regardless of renal function. |
| 28 | Koretsune et. Al. 2015 | Japan | 93 | 60 | 33 | Edoxaban | 50 | Warfarin | 43 | High | The study found that **edoxaban 15 mg once daily** is **safe and effective** for **Japanese patients with severe renal impairment and non-valvular atrial fibrillation**, showing similar outcomes to higher doses in patients with normal or mild renal impairment. |
| 29 | Spencer et. Al. 2015 | Multicenter | 14264 | 8558 | 5706 | Rivaroxaban | 7111 | Warfarin | 7125 | Moderate | The main finding of the ROCKET AF trial is that rivaroxaban is non-inferior to warfarin for preventing stroke and systemic embolism in patients with non-valvular atrial fibrillation, with similar bleeding rates but lower intracranial bleeding. |
| 30 | Bohula et. Al. 2016 | Multicenter | 21105 | 9709 | 11396 | Edoxaban | 10535 | Warfarin | 10570 | High | The main finding of the study is that **higher-dose edoxaban (HDER) demonstrated comparable efficacy to warfarin for preventing thromboembolic events in atrial fibrillation patients, with significantly lower rates of major bleeding**. |
| 31 | Alexander et. Al. 2018 | Multicenter | 16800 | 8400 | 8400 | Apixaban | 9120 | Warfarin | 9081 | High | The main finding of the study is that **apixaban** is more effective and safer than warfarin in patients with atrial fibrillation and multi-morbidity, with preserved efficacy and reduced bleeding risks across different multi-morbidity groups. |
| 32 | Lullo et. Al. 2018 | Italy | 347 | 192 | 155 | Rivaroxaban | 247 | Warfarin | 100 | Moderate | The main finding of the study is that rivaroxaban appears to be a safe and effective therapeutic option for patients with moderate-to-advanced chronic kidney disease, showing a better risk-to-benefit profile compared to warfarin. |
| 33 | Coleman et. Al. 2019 | USA | 6638 | 4613 | 2025 | Rivaroxaban | 3319 | Warfarin | 3319 | High | The main finding of the study is that rivaroxaban significantly reduced the rate of stroke or systemic embolism compared to warfarin in patients with non-valvular atrial fibrillation and a CHA2DS2-VASc score of 1, without increasing major bleeding events. |
| 34 | Schafer et. Al. 2018 | USA | 604 | 302 | 302 | Apixaban | 302 | Warfarin | 302 | High | The study found that apixaban had similar or lower major bleeding rates compared to warfarin in patients with advanced chronic kidney disease, with no significant differences in stroke or thromboembolism rates. |
| 35 | Hernandez et. Al. 2019 | USA | 21682 | 13812 | 7870 | Rivaroxaban | 10017 | Warfarin | 11665 | High | The study found that rivaroxaban was associated with a lower risk of acute kidney injury and progression to Stage 5 chronic kidney disease or need for dialysis compared to warfarin in diabetic patients with non-valvular atrial fibrillation. |
| 36 | Tanaka et. Al. 2020 | Japan | 51 | 49 | 4 | Rivaroxaban | 25 | Warfarin | 26 | Moderate | The main finding of the study is that rivaroxaban, compared to warfarin, had neutral effects on urinary albumin excretion in patients with atrial fibrillation and chronic kidney disease over 3 months. |
| 37 | Vriese et. Al. 2021 | Belgium | 132 | 88 | 44 | Rivaroxaban | 46 | Warfarin | 44 | High | The main finding of the study is that rivaroxaban significantly reduced the composite outcome of fatal and nonfatal cardiovascular events and major bleeding complications compared with vitamin K antagonists (VKAs) in hemodialysis patients with atrial fibrillation. |
| 39 | Cho et. Al. 2022 | South Korea | 150 | 75 | 75 | Rivaroxaban | 75 | Warfarin | 75 | Moderate | The main finding of the study is that rivaroxaban may reduce myocardial and renal injury in patients with acute decompensated heart failure and atrial fibrillation, compared to warfarin, based on biomarker levels. |
| 40 | Fordyce et. Al. 2016 | Multicenter | 12612 | 7653 | 4959 | Rivaroxaban | 6359 | Warfarin | 6253 | High | The main finding is that among patients with worsening renal function, rivaroxaban was associated with lower rates of stroke and systemic embolism compared to warfarin, without an increase in major or non-major clinically relevant bleeding events. |
| 41 | Hijazi et. Al. 2021 | Multicenter | 4456 | 3155 | 1301 | Apixaban | 2228 | Aspirin | 2228 | High | The main finding of the study is that apixaban, compared to vitamin K antagonists, consistently showed lower rates of bleeding and hospitalization, with no significant difference in ischemic events across various kidney function categories. |
| 42 | Reinecke et. Al. 2023 | Germany | 97 | 68 | 29 | Apixaban | 48 | Aspirin | 49 | Moderate | The main finding of the study is that apixaban (2.5 mg BID) showed no significant difference in safety or efficacy compared to phenprocoumon (VKA) in patients with atrial fibrillation on chronic hemodialysis. |
| 43 | Novikova et. Al. 2021 | Russia | 133 | 29 | 50 | Dabigatran | 79 | Warfarin | 54 | High | The main finding of the study is that dabigatran therapy in patients with atrial fibrillation and chronic kidney disease leads to a moderate decline in glomerular filtration rate (GFR) over five years, with no significant worsening in patients with initially low GFR. |
| 44 | Nash et. Al. 2025 | UK | 3648 | 2008 | 3648 | Apixaban | 3058 | Warfarin | 543 | High | The main finding of the OPTIMAS trial is that early initiation of direct oral anticoagulants (DOACs) after acute ischemic stroke in patients with atrial fibrillation and chronic kidney disease is safe and noninferior to delayed initiation, without increasing bleeding risk. |
| 45 | OCEANIC-AF et. Al. 2025 | Multicenter | 14810 | 9596 | 5214 | Asundexian | 7415 | Apixaban | 7395 | Moderate | The main finding of the trial is that asundexian, a factor XIa inhibitor, was associated with a higher incidence of stroke or systemic embolism but fewer major bleeding events compared to apixaban in patients with atrial fibrillation at risk for stroke. |
| 46 | Suzuki et. Al. 2015 | Japan | 160 | 80 | 80 | Rivaroxaban | 80 | Warfarin | 80 | High | The main finding of the study is that the X-NOAC trial is evaluating the efficacy of rivaroxaban compared to warfarin on renal function in patients with non-valvular atrial fibrillation and chronic kidney disease. |

Table S2. Relative treatment effects and ranking for all studies in overall survival

|  | **Apixaban** | **Aspirin** | **Asundexian** | **Dabigatran** | **Edoxaban** | **Rivaroxaban** |
| --- | --- | --- | --- | --- | --- | --- |
| Apixaban | Apixaban | 0.8 (0.26, 2.46) | 0.06 (0.01, 0.36) | 0.34 (0.07, 1.75) | 0.22 (0.04, 0.97) | 1.15 (0.45, 2.99) |
| Aspirin | 1.26 (0.41, 3.86) | Aspirin | 0.08 (0.01, 0.63) | 0.42 (0.06, 3.07) | 0.27 (0.04, 1.76) | 1.45 (0.33, 6.3) |
| Asundexian | 15.89 (2.81, 76.74) | 12.67 (1.58, 84.67) | Asundexian | 5.39 (0.47, 49.12) | 3.48 (0.31, 30.02) | 18.37 (2.49, 114.09) |
| Dabigatran | 2.97 (0.57, 15.25) | 2.37 (0.33, 17.29) | 0.19 (0.02, 2.11) | Dabigatran | 0.65 (0.08, 4.58) | 3.44 (0.69, 16.77) |
| Edoxaban | 4.57 (1.03, 22.32) | 3.64 (0.57, 26.17) | 0.29 (0.03, 3.23) | 1.54 (0.22, 11.94) | Edoxaban | 5.29 (1.25, 24.55) |
| Rivaroxaban | 0.87 (0.33, 2.24) | 0.69 (0.16, 3.05) | 0.05 (0.01, 0.4) | 0.29 (0.06, 1.45) | 0.19 (0.04, 0.8) | Rivaroxaban |

Table S3. Sensitivity Analysis of Relative treatment effects and ranking for all studies in overall survival

|  | **Apixaban** | **Aspirin** | **Asundexian** | **Dabigatran** | **Edoxaban** | **Rivaroxaban** |
| --- | --- | --- | --- | --- | --- | --- |
| Apixaban | Apixaban | 0.94 (0.26, 3.3) | 0.02 (0, 0.12) | 0.33 (0.05, 2.2) | 0.43 (0.08, 2.26) | 1.15 (0.46, 2.9) |
| Aspirin | 1.06 (0.3, 3.8) | Aspirin | 0.02 (0, 0.2) | 0.35 (0.04, 3.45) | 0.46 (0.06, 3.72) | 1.22 (0.26, 5.95) |
| Asundexian | 45.4 (8.55, 240.61) | 42.67 (5.12, 349.08) | Asundexian | 14.87 (1.18, 187.96) | 19.42 (1.77, 200.39) | 52.39 (7.64, 352.5) |
| Dabigatran | 3.05 (0.45, 20.35) | 2.87 (0.29, 27.68) | 0.07 (0.01, 0.85) | Dabigatran | 1.31 (0.12, 13.19) | 3.5 (0.54, 23.08) |
| Edoxaban | 2.33 (0.44, 12.69) | 2.19 (0.27, 18.11) | 0.05 (0, 0.57) | 0.76 (0.08, 8.19) | Edoxaban | 2.69 (0.54, 14) |
| Rivaroxaban | 0.87 (0.34, 2.18) | 0.82 (0.17, 3.84) | 0.02 (0, 0.13) | 0.29 (0.04, 1.84) | 0.37 (0.07, 1.86) | Rivaroxaban |

Table S4. Assessment of Inconsistency in all studies for overall survival

| **Comparison** | **No.Studies** | **NMA** | **Direct** | **Indirect** | **Difference** | **Diff_95CI_lower** | **Diff_95CI_upper** | **pValue** |
| --- | --- | --- | --- | --- | --- | --- | --- | --- |
| Apixaban:Aspirin | 3 | 0.22 | 0.22 | NA | NA | NA | NA | NA |
| Apixaban:Asundexian | 2 | 2.79 | 2.79 | NA | NA | NA | NA | NA |
| Apixaban:Dabigatran | 0 | 1.07 | NA | 1.07 | NA | NA | NA | NA |
| Apixaban:Edoxaban | 0 | 1.29 | NA | 1.29 | NA | NA | NA | NA |
| Apixaban:Rivaroxaban | 0 | -0.14 | NA | -0.14 | NA | NA | NA | NA |
| Apixaban:Warfarin | 7 | 0.69 | 0.69 | NA | NA | NA | NA | NA |
| Aspirin:Asundexian | 0 | 2.57 | NA | 2.57 | NA | NA | NA | NA |
| Aspirin:Dabigatran | 0 | 0.85 | NA | 0.85 | NA | NA | NA | NA |
| Aspirin:Edoxaban | 0 | 1.07 | NA | 1.07 | NA | NA | NA | NA |
| Aspirin:Rivaroxaban | 0 | -0.36 | NA | -0.36 | NA | NA | NA | NA |
| Aspirin:Warfarin | 0 | 0.47 | NA | 0.47 | NA | NA | NA | NA |
| Asundexian:Dabigatran | 0 | -1.72 | NA | -1.72 | NA | NA | NA | NA |
| Asundexian:Edoxaban | 0 | -1.50 | NA | -1.50 | NA | NA | NA | NA |
| Asundexian:Rivaroxaban | 0 | -2.93 | NA | -2.93 | NA | NA | NA | NA |
| Asundexian:Warfarin | 0 | -2.10 | NA | -2.10 | NA | NA | NA | NA |
| Dabigatran:Edoxaban | 0 | 0.22 | NA | 0.22 | NA | NA | NA | NA |
| Dabigatran:Rivaroxaban | 0 | -1.21 | NA | -1.21 | NA | NA | NA | NA |
| Dabigatran:Warfarin | 2 | -0.38 | -0.38 | NA | NA | NA | NA | NA |
| Edoxaban:Rivaroxaban | 0 | -1.43 | NA | -1.43 | NA | NA | NA | NA |
| Edoxaban:Warfarin | 3 | -0.60 | -0.60 | NA | NA | NA | NA | NA |
| Rivaroxaban:Warfarin | 11 | 0.83 | 0.83 | NA | NA | NA | NA | NA |

Table S5. Sensitivity analysis of all the studies included for overall survival

| **Comparison** | **No.Studies** | **NMA** | **Direct** | **Indirect** | **Difference** | **Diff_95CI_lower** | **Diff_95CI_upper** | **pValue** |
| --- | --- | --- | --- | --- | --- | --- | --- | --- |
| Apixaban:Aspirin | 2 | 0.05 | 0.05 | NA | NA | NA | NA | NA |
| Apixaban:Asundexian | 1 | 3.81 | 3.81 | NA | NA | NA | NA | NA |
| Apixaban:Dabigatran | 0 | 1.13 | NA | 1.13 | NA | NA | NA | NA |
| Apixaban:Edoxaban | 0 | 0.80 | NA | 0.80 | NA | NA | NA | NA |
| Apixaban:Rivaroxaban | 0 | -0.13 | NA | -0.13 | NA | NA | NA | NA |
| Apixaban:Warfarin | 6 | 0.81 | 0.81 | NA | NA | NA | NA | NA |
| Aspirin:Asundexian | 0 | 3.76 | NA | 3.76 | NA | NA | NA | NA |
| Aspirin:Dabigatran | 0 | 1.08 | NA | 1.08 | NA | NA | NA | NA |
| Aspirin:Edoxaban | 0 | 0.75 | NA | 0.75 | NA | NA | NA | NA |
| Aspirin:Rivaroxaban | 0 | -0.18 | NA | -0.18 | NA | NA | NA | NA |
| Aspirin:Warfarin | 0 | 0.76 | NA | 0.76 | NA | NA | NA | NA |
| Asundexian:Dabigatran | 0 | -2.68 | NA | -2.68 | NA | NA | NA | NA |
| Asundexian:Edoxaban | 0 | -3.01 | NA | -3.01 | NA | NA | NA | NA |
| Asundexian:Rivaroxaban | 0 | -3.94 | NA | -3.94 | NA | NA | NA | NA |
| Asundexian:Warfarin | 0 | -3.00 | NA | -3.00 | NA | NA | NA | NA |
| Dabigatran:Edoxaban | 0 | -0.33 | NA | -0.33 | NA | NA | NA | NA |
| Dabigatran:Rivaroxaban | 0 | -1.26 | NA | -1.26 | NA | NA | NA | NA |
| Dabigatran:Warfarin | 1 | -0.32 | -0.32 | NA | NA | NA | NA | NA |
| Edoxaban:Rivaroxaban | 0 | -0.93 | NA | -0.93 | NA | NA | NA | NA |
| Edoxaban:Warfarin | 2 | 0.01 | 0.01 | NA | NA | NA | NA | NA |
| Rivaroxaban:Warfarin | 10 | 0.94 | 0.94 | NA | NA | NA | NA | NA |

Figure S1. Funnel Plots of Thrombotic events

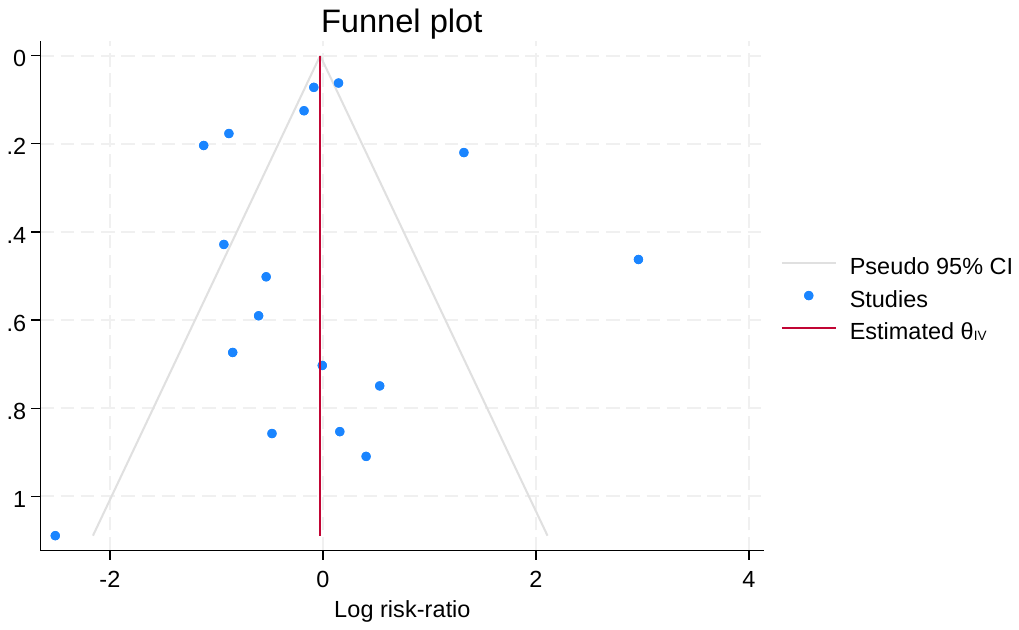

Table S6. Relative treatment effects and ranking for all studies in Thrombotic Events

|  | **Apixaban** | **Aspirin** | **Asundexian** | **Dabigatran** | **Edoxaban** | **Rivaroxaban** | **Warfarin** |
| --- | --- | --- | --- | --- | --- | --- | --- |
| Apixaban | Apixaban | 2.61 (0.49, 13.42) | 2.45 (0.39, 12.78) | 2.36 (0.11, 49.63) | 3.68 (0.51, 78.84) | 0.64 (0.11, 3.12) | 1.41 (0.36, 5.26) |
| Aspirin | 0.38 (0.07, 2.02) | Aspirin | 0.94 (0.08, 9.47) | 0.9 (0.03, 28.3) | 1.4 (0.12, 52.03) | 0.25 (0.02, 2.32) | 0.54 (0.06, 4.49) |
| Asundexian | 0.41 (0.08, 2.54) | 1.06 (0.11, 12.76) | Asundexian | 0.97 (0.03, 34.33) | 1.5 (0.13, 62.61) | 0.26 (0.02, 2.92) | 0.58 (0.07, 5.61) |
| Dabigatran | 0.42 (0.02, 9.34) | 1.11 (0.04, 35.34) | 1.03 (0.03, 32.59) | Dabigatran | 1.59 (0.08, 86.84) | 0.27 (0.01, 4.89) | 0.6 (0.04, 9.51) |
| Edoxaban | 0.27 (0.01, 1.96) | 0.72 (0.02, 8.11) | 0.67 (0.02, 7.81) | 0.63 (0.01, 12.19) | Edoxaban | 0.17 (0.01, 1.03) | 0.38 (0.03, 1.83) |
| Rivaroxaban | 1.56 (0.32, 9.32) | 4.06 (0.43, 47.44) | 3.81 (0.34, 43.75) | 3.69 (0.2, 73.29) | 5.77 (0.97, 124.68) | Rivaroxaban | 2.19 (0.86, 6.7) |
| Warfarin | 0.71 (0.19, 2.77) | 1.85 (0.22, 15.51) | 1.73 (0.18, 14.77) | 1.68 (0.11, 25.74) | 2.6 (0.55, 39.6) | 0.46 (0.15, 1.16) | Warfarin |

Table S7. Relative treatment effects and ranking for all studies in Thrombotic Events for sensitivity analysis

|  | **Apixaban** | **Aspirin** | **Asundexian** | **Dabigatran** | **Edoxaban** | **Rivaroxaban** | **Warfarin** |
| --- | --- | --- | --- | --- | --- | --- | --- |
| Apixaban | Apixaban | 2.65 (0.5, 13.67) | 2.5 (0.4, 12.64) | 1.94 (0.08, 40.82) | 2.76 (0.37, 80.45) | 0.64 (0.09, 3.98) | 1.14 (0.23, 5.29) |
| Aspirin | 0.38 (0.07, 2.02) | Aspirin | 0.95 (0.08, 9.39) | 0.73 (0.02, 22.93) | 1.03 (0.09, 49.62) | 0.24 (0.02, 2.78) | 0.43 (0.04, 4.04) |
| Asundexian | 0.4 (0.08, 2.52) | 1.06 (0.11, 12.81) | Asundexian | 0.78 (0.02, 28.19) | 1.1 (0.09, 62.42) | 0.26 (0.02, 3.44) | 0.46 (0.05, 5) |
| Dabigatran | 0.51 (0.02, 13.06) | 1.36 (0.04, 49.75) | 1.28 (0.04, 46.86) | Dabigatran | 1.46 (0.08, 98.33) | 0.33 (0.02, 6.15) | 0.59 (0.04, 9.45) |
| Edoxaban | 0.36 (0.01, 2.71) | 0.97 (0.02, 11.13) | 0.91 (0.02, 10.82) | 0.69 (0.01, 12.45) | Edoxaban | 0.23 (0.01, 1.31) | 0.42 (0.02, 1.85) |
| Rivaroxaban | 1.55 (0.25, 11.51) | 4.11 (0.36, 56.49) | 3.86 (0.29, 50.39) | 3.02 (0.16, 58.3) | 4.26 (0.76, 107.86) | Rivaroxaban | 1.77 (0.63, 5.63) |
| Warfarin | 0.88 (0.19, 4.35) | 2.31 (0.25, 23.24) | 2.17 (0.2, 21.32) | 1.69 (0.11, 24.66) | 2.38 (0.54, 44.16) | 0.57 (0.18, 1.6) | Warfarin |

Figure S2. Funnel Plot of Bleeding Events

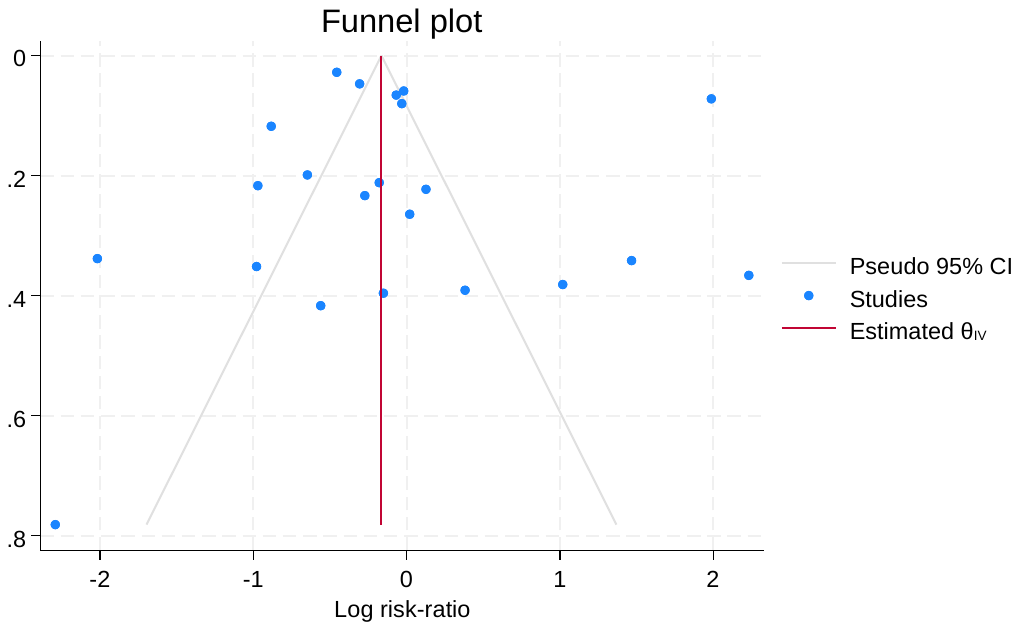

Table S9. Relative treatment effects and ranking for all studies in Bleeding Events

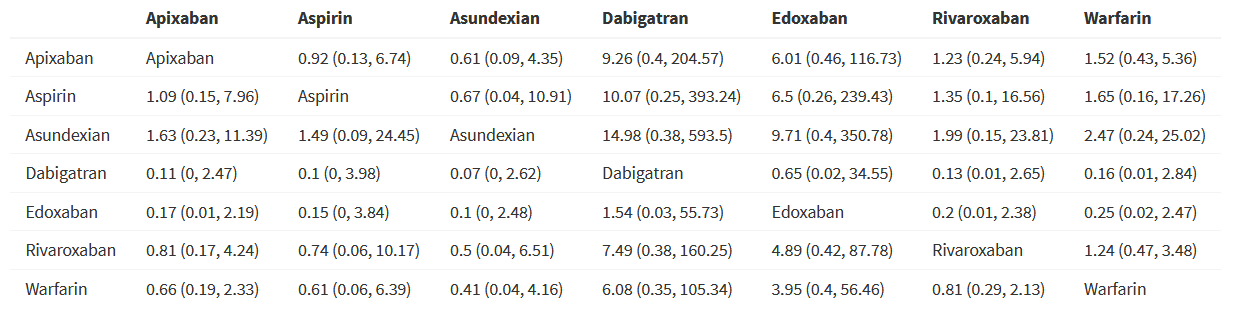

Figure S3. Funnel Plot of All Cause Mortality

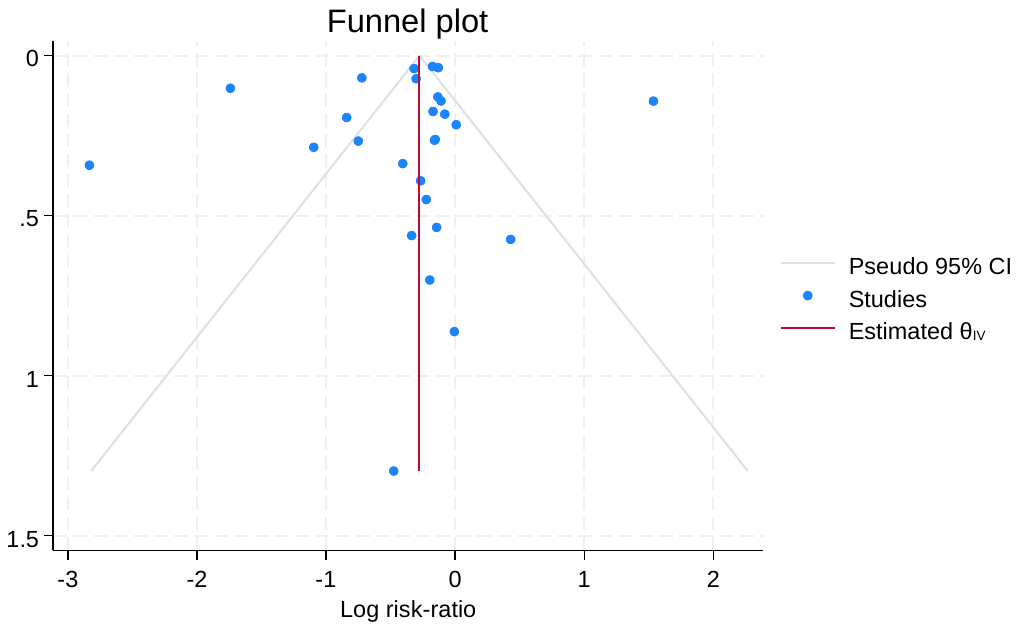

Figure S4. ROBS 2.0

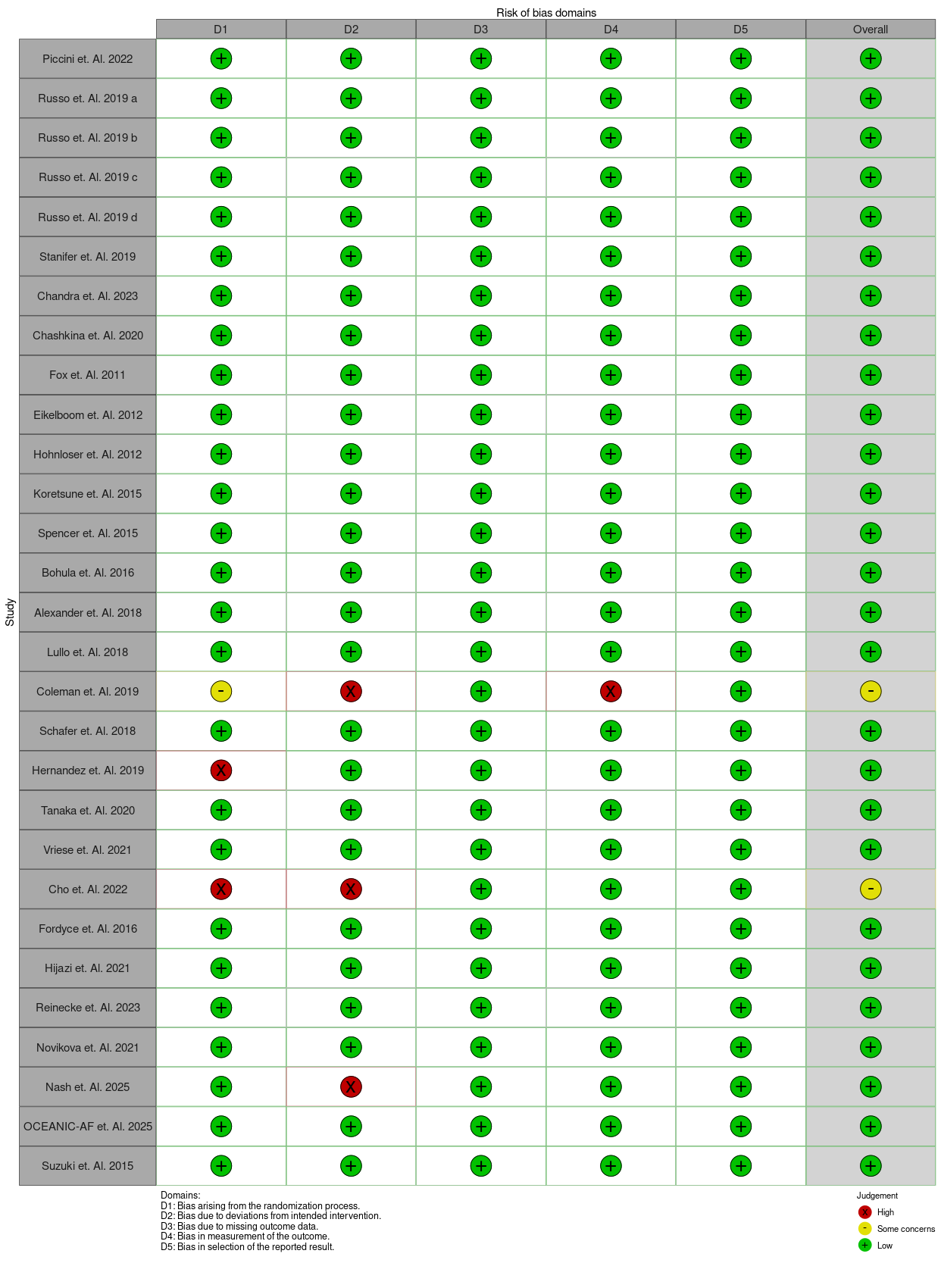
